# Multisystemic inflammatory syndrome following COVID-19 mRNA vaccine in children: a national post-authorization pharmacovigilance study

**DOI:** 10.1101/2022.01.17.22269263

**Authors:** Naïm Ouldali, Haleh Bagheri, Francesco Salvo, Denise Antona, Antoine Pariente, Claire Leblanc, Martine Tebacher, Joëlle Micallef, Corinne Levy, Robert Cohen, Etienne Javouhey, Brigitte Bader-Meunier, Caroline Ovaert, Sylvain Renolleau, Veronique Hentgen, Isabelle Kone-Paut, Nina Deschamps, Loïc De Pontual, Xavier Iriart, Christelle Gras-Le Guen, François Angoulvant, Alexandre Belot, the “French Covid-19 Paediatric Inflammation Consortium”, the “French Pharmacovigilance network”

## Abstract

**Importance:** Multisystem inflammatory syndrome in children (MIS-C) is the most severe life-threatening clinical entity associated with pediatric SARS-CoV-2 infection. Whether COVID-19 mRNA vaccine can induce this complication in children is unknown.

**Objective:** To assess the risk of hyper-inflammatory syndrome following COVID-19 mRNA vaccine in children.

**Design, Setting, and Participants:** Post-authorization national population-based surveillance using the French enhanced pharmacovigilance surveillance system for COVID-19 vaccines. All cases of suspected hyper-inflammatory syndrome following COVID-19 mRNA vaccine in 12– 17-year-old children between June 15^th^, 2021 and January 1^st^, 2022, were reported. Each case was assessed for WHO MIS-C criteria. Causality assessment followed 2019 WHO recommendations.

**Exposure:** COVID-19 mRNA vaccine.

**Main Outcome and Measures:** The main outcome was the reporting rate of post-vaccine hyper-inflammatory syndrome per 1,000,000 COVID-19 mRNA vaccine doses in 12–17-year-old children. This reporting rate was compared to the MIS-C rate per 1,000,000 12–17-year-old children infected by SARS-CoV-2. Secondary outcomes included the comparison of clinical features between post-vaccine hyper-inflammatory syndrome and post SARS-CoV-2 MIS-C.

**Results:** From June 2021 to January 2022, 8,113,058 COVID-19 mRNA vaccine doses were administered to 4,079,234 12–17-year-old children. Among them, 9 presented a multisystemic hyper-inflammatory syndrome. All cases fulfilled MIS-C WHO criteria. Main clinical features included male predominance (8/9, 89%), cardiac involvement (8/9, 89%), digestive symptoms (7/9, 78%), coagulopathy (5/9, 54%), cytolytic hepatitis (4/9, 46%), and shock (3/9, 33%). 3/9 (33%) required intensive care unit transfer, and 2/9 (22%) hemodynamic support. All cases recovered. Only three cases had evidence of previous SARS-CoV-2 infection. The reporting rate was 1.1 (95%CI [0.5; 2.1]) per 1,000,000 doses injected. As a comparison, 113 MIS-C (95%CI [95; 135]) occurred per 1,000,000 12–17-year-old children infected by SARS-CoV-2. Clinical features (inflammatory parameters, cytopenia) slightly differed from post-SARS-CoV-2 MIS-C, along with short-term outcomes (less PICU transfer than MIS-C).

**Conclusion and Relevance:** Very few cases of hyper-inflammatory syndromes with multi-organ involvement occurred following COVID-19 mRNA vaccine in 12–17-year-old children. The low reporting rate of this syndrome, compared to the rate of MIS-C among same age children infected by SARS-CoV-2, supports the benefit of SARS-CoV-2 vaccination in children. Further studies are required to explore specific pathways of this entity compared to post-SARS-CoV-2 MIS-C.

**Key points:** *Question:* Is COVID-19 mRNA vaccine in 12-17-year-old children associated with subsequent multisystemic hyper-inflammatory syndrome?

*Findings:* The French national pharmacovigilance system identified 9 children with a hyper-inflammatory syndrome with multi-organ involvement following COVID-19 mRNA vaccination (reporting rate 1.1 [0.5; 2.1] per 1,000,000 doses), of which only three had evidence of previous SARS-CoV-2 infection. All cases fulfilled WHO definition for MIS-C, but clinical and immunological features, along with short-term outcomes, slightly differed from classical post SARS-CoV-2 MIS-C.

*Meaning:* Very rare cases of hyper-inflammatory syndrome can occur following COVID-19 mRNA vaccine in 12-17-year-old children. The very low rate of this entity, compared to classical post-SARS-CoV-2 MIS-C, supports the benefit of SARS-CoV-2 vaccination in children.

## Background

Multisystem inflammatory syndrome in children (MIS-C) is a novel clinical entity first described in April 2020.^1–5^ Its association with SARS-CoV-2 infection has been documented, with a previous infection occurring 4 to 6 weeks before MIS-C onset.^4–6^ The main clinical features of MIS-C are frequent acute cardiac dysfunction, shock, multi organ failure that often require pediatric intensive care unit transfer and hemodynamic support.^7^ Thus, numerous studies showed that MIS-C is by far the most severe form associated with SARS-CoV-2 infection in children and the leading source of morbidity related to SARS-CoV-2 in this age group.^7,8^

The pathophysiology of this disease remains unknown, but previous investigations showed that MIS-C is characterized by a cytokine storm^9^ associated with a superantigen-like activation of T cells with an expansion of Vβ21.3-expressing T cells which is not seen in toxic shock syndrome, Kawasaki disease or other COVID-19 features^10–13^. Notably, SARS-CoV2 spike harbors a motif located in the receptor binding domain the that is predicted in silico to interact with Vβ region in T cells. Whether antigenic exposure limited to the Spike protein can lead to similar dysregulated immune response remains unknown.

Two COVID-19 mRNA vaccines have been shown to be efficacious and well tolerated in adults, and have been introduced since December 2020.^14^ Post-authorization studies confirmed their major impact on SARS-CoV-2 epidemics,^15^ with very few serious adverse events reported to date.^16,17^ In children, the immunogenicity, efficacy, as well as frequent adverse events have been assessed in trials involving thousands of 12-17-year-old children.^18^ Based on these studies, the Food and Drug Administration (FDA) and European Medicines Agency (EMA) authorized formulations of BNT162b2 COVID-19 vaccine for ages 12-17-year. However, rare serious adverse events following immunization could not be detected by these clinical trials. Especially, whether exposure to SARS-CoV-2 antigens due to mRNA vaccine can induce MIS-C is unknown.

Given the lower burden of SARS-CoV-2 related diseases in children compared to adults, elucidating the safety profile of mRNA vaccine, especially regarding MIS-C, is of critical interest to establish its benefit-risk balance in this population. In this context, monitoring post-vaccine MIS-C has been identified as a priority by the FDA, the EMA, and the French National Agency for Medicines and Health Products safety (ANSM).^19–21^ Several cases reports of children with MIS-C following mRNA vaccine recently raised major concerns regarding this potential vaccine-related adverse event.^22–25^

Using a well-established national pharmacovigilance surveillance system coordinated by ANSM,^26,27^ we aimed to evaluate the potential association of COVID-19 mRNA vaccine and subsequent hyper-inflammatory syndrome in children.

## Methods

### Ethical review

For the pharmacovigilance surveillance system, this study was performed according to the authorization from the National Commission on Informatics and Liberty (CNIL) n° 2014-302 for the national pharmacovigilance database done by ANSM. For the MIS-C following SARS-CoV-2 infection surveillance system, the study was approved by the INSERM ethics committee for evaluation (IRB00003888). A written information form validated by the ethics committee was given to all participants. Oral consent was obtained from study participants; no family members or participants refused to participate.

### Study design and settings

We conducted a post-authorization prospective national population-based surveillance using the well-established ANSM pharmacovigilance system.^26,27^ This network is based on 31 regional pharmacovigilance centers, which cover all the French territory, and is coordinated by ANSM since 1973.^26^ All ambulatory or hospital-based health practitioners throughout France or patient that observe a suspected adverse drug reaction (ADR) report the event to the regional center via a secure platform.^26^ Reporting of all ADRs, independently of the seriousness or expectedness, is compulsory for health practitioners. All reports undergo a pharmacological, clinical and biological assessment process by a trained assessor of the Regional Center. Cases are then registered in the national computerized pharmacovigilance database, centralized at ANSM, to allow anonymized case reviewing at a national level by ANSM and experts in the field, drug causality assessment, and to recommend specific measures if required.^27^ The detailed methodology of this French pharmacovigilance system has been previously published.^26,27^

As part of the national COVID-19 vaccination campaign, ANSM had put in place a specific reinforced surveillance system to provide real-time monitoring of COVID-19 vaccines ADRs at a national level.^21,28^ This is part of the risk management plan coordinated by the European Medicines Agency (EMA). The objectives are to carry out a continuous assessment of the safety of vaccines COVID-19 vaccines in order to confirm their safety or to quickly take the relevant measures, and to allow the Health Ministry to adapt the vaccination strategy, if necessary. For each marketed COVID-19 vaccine, two to five regional pharmacovigilance centers have been designated to gather and assess on a daily basis all adverse drug reactions reported following immunization. An expert of the organ involved is solicited to analyze the reported cases every week, to identify atypical and/or serious patterns leading to potential safety signals.^21^ Then a weekly meeting involving ANSM and all regional pharmacovigilance centers is organized to discuss the expert pharmacovigilance reports, potential safety signals, and new data from the literature, in order to confirm or not safety signals.^21,28^ A complete report including the synopsis of these meeting are published by ANSM every two weeks.^21,28^ If a national safety signal is validated, appropriate measures are issued in relation with European Medicine Agency to prevent or reduce the likelihood of the risk occurring in vaccinated people. The detailed methodology of this specific COVID-19 vaccine monitoring is available elsewhere.^21,28^

### Cases review to assess WHO criteria for MIS-C and vaccine causality

All pediatric cases of inflammatory syndrome, fever > 3 days, shock, or acute organ dysfunction without any obvious cause, occurring any time after COVID-19 mRNA vaccine injection in children under 18 years of age in France from June 15^th^, 2021, to January 1^st^, 2022, were eligible. Following 2019 WHO guidelines for causality assessment of an adverse event following immunization,^29^ each case was reviewed by a multidisciplinary committee, with experts in pediatric immunization, pediatric infectious diseases, pediatric rheumatology, immunology and internal medicine, pediatric intensive care, pediatric cardiology, and experts pharmacologists from pharmacovigilance centers. All these experts were involved in MIS-C surveillance and management in France as part of the French MIS-C consortium, and developed specific expertise in this field.^5,30,31^ Medical records were obtained for all cases to accurately assess if cases fulfilled WHO criteria for MIS-C. Cases were included after reviewing if they fulfilled WHO MIS-C criteria, with a delay between the last vaccine administration and disease onset < 2 months, based on available data from the literature regarding the delay between SARS-CoV-2 infection and MIS-C onset.^4–6^ An important part of the vaccine causality assessment relied on identifying other potential causes for the event.^29^ For hyper-inflammatory syndrome, extensive investigation of previous exposure to SARS-CoV-2 over the past two months was critical, and relied on investigating history of documented infection, and performing nasopharyngeal SARS-CoV-2 Polymerase chain reaction (PCR) and anti-Nucleocapsid (anti-N) serology.^32^

### National immunization program

BNT162b2 COVID-19 mRNA vaccine have been introduced for 12-17-year-old children on June 15^th^, 2021 in France.^33^ It has been followed by mRNA-1273 approval for same age children on July, 27^th^, 2021.^33^ A higher risk of myocarditis or pericarditis has been suggested following mRNA-1273 compared to BNT162b2 in adults younger than 30 years old.^34^ This has led French authorities to prioritize BNT162b2 for 12-17-year-old children immunization. Thus, by January 1^st^, 2022, the large majority of vaccinated 12-17-year-old children received BNT162b2 (>95%).^31^

### Outcome measure

The main outcome was the national reporting rate of hyper-inflammatory syndrome following COVID-19 mRNA vaccine per 1,000,000 doses in 12-17-year-old children in France. To calculate national reporting rate, we used as a denominator the total number of COVID-19 mRNA vaccine dose administered in 12-17-year-old children over the study period (available at https://solidarites-sante.gouv.fr/grands-dossiers/vaccin-covid-19/article/le-tableau-de-bord-de-la-vaccination). We also estimated in the same age-group, in the same population, the rate of post-SARS-CoV-2 MIS-C cases per 1,000,000 infections in 12-17-year-old children in France for comparison.

Secondary outcomes were the reporting rate of hyper-inflammatory syndrome following first and second injections of COVID-19 mRNA vaccine in 12-17-year-old children in France, reporting rate by sex, and comparison of clinical features of hyper-inflammatory syndrome following COVID-19 mRNA vaccine versus post-SARS-CoV-2 MIS-C cases.

### MIS-C following SARS-CoV-2 infection surveillance system

To estimate the rate of post-SARS-CoV-2 MIS-C cases per 1,000,000 infections in 12-17-year-old children in France, we used data from the French COVID-19 Pediatric Inflammation Consortium, coordinated by Public health France.^5,30,31^ As previously published, since April 2020, all suspected MIS-C cases in France were mandatorily reported to Public health France. Each suspected case was then assessed following WHO criteria for MIS-C.^5,30,31^ Furthermore, Public health France also conducted seroprevalence studies that allowed estimating the proportion of 12-17-year-old old children infected by SARS-CoV-2 since the beginning of the pandemic.^35^ Thus, to estimate the rate of post-SARS-CoV-2 MIS-C cases per 1,000,000 infections, we used as a numerator the number of confirmed 12-17-year-old MIS-C cases reported to Public health France since the start of the pandemic, and as a denominator the estimated number of 12-17-year-old French children infected by SARS-CoV-2 since the start of the pandemic. To avoid any bias in MIS-C rate estimation due to vaccine implementation,^31^ we restricted this analysis to the pre-vaccine period, i.e. from the start of the pandemic to June 15^th^, 2021.

This surveillance system also collected clinico-biological and short term outcome data of MIS-C cases that fulfilled WHO criteria.^5,30,31^ Thus, to compare the clinical presentation of hyper-inflammatory syndrome following COVID-19 mRNA vaccine to post-SARS-CoV-2 MIS-C cases in the same population, we included all unvaccinated 12-17-year-old MIS-C cases fulfilling WHO criteria with available clinical files as same-age and same-population comparator group.

### Statistical analysis

We describe patient characteristics with numbers (percentages) for categorical variables and median (interquartile range [IQR]) for quantitative variables. We compared clinical and biological characteristics using non-parametric Fisher’s exact test for categorical variables and Mann-Whitney U test for quantitative variables. A two-sided p-value <0.05 was considered statistically significant. Reporting rate of hyper-inflammatory syndrome was expressed as cases per 1,000,000 vaccine injections with 95% CIs. All statistical analyses involved using R v3.6.1 (http://www.R-project.org).

## Results

From June 15^th^, 2021 to January 1^st^, 2022, 8,113,058 COVID-19 mRNA vaccine doses were administered to 4,079,234 12-17-year-old children in France (including 4,079,234 first injections, 3,905,636 second injections, and 128,188 third injections). Over this period, 2,028 adverse drug reactions related to COVID-19 mRNA vaccine have been reported to the pharmacovigilance centers in 12-17-year-old children. Among them, 9 cases of hyper-inflammatory syndrome were reported. All cases fulfilled WHO criteria for MIS-C (Table 1). All cases involved BNT162b2 vaccine (5 cases following the first injection, 4 following the second injection). The delay between last injection and disease onset ranged from 2 days to 42 days.

**Table 1.**
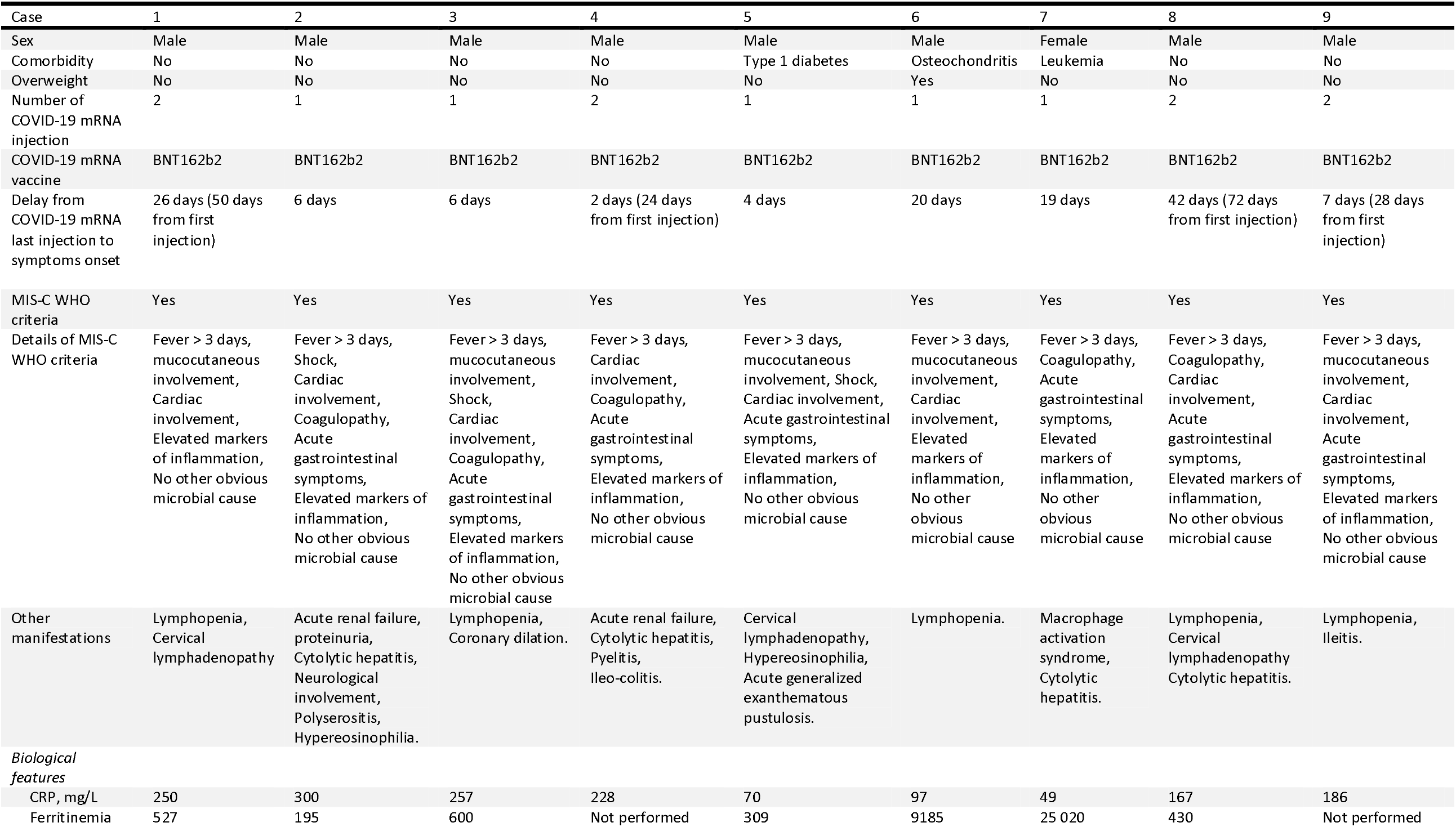
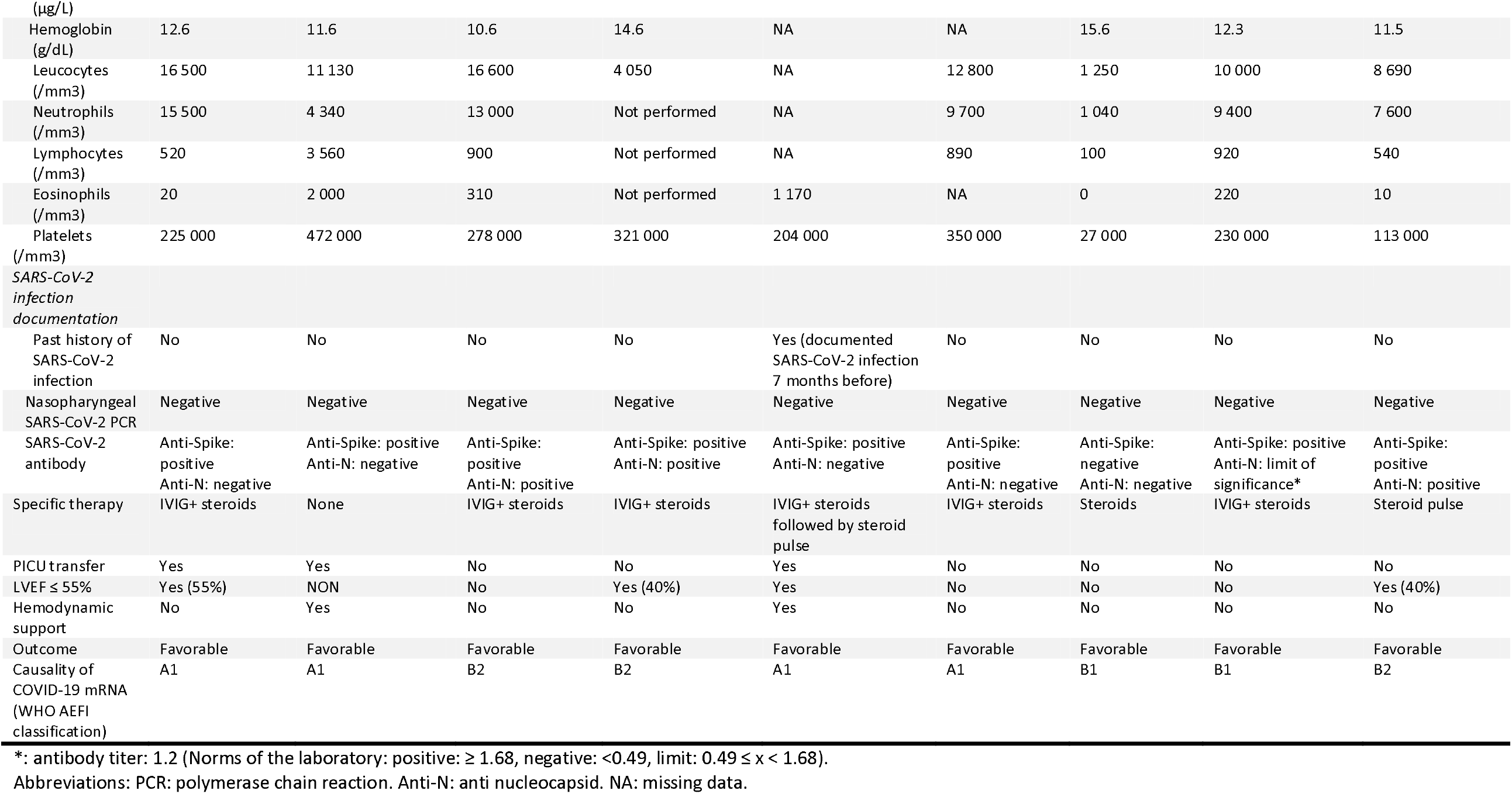
Characteristics of children with hyper inflammatory syndrome following COVID-19 mRNA in France.

### Investigation of previous SARS-CoV-2 infection

All 9 cases had complete data for history of documented infection, nasopharyngeal SARS-CoV-2 PCR, and anti-N serology. In one case, a previous infection 7 months before disease onset was reported, but was too far apart to be considered as linked to the disease. All children had negative SARS-CoV-2 PCR, and three children had a positive anti-N serology (Table 1). Based on all information available, the mRNA vaccine causality was considered consistent in 4 cases, and indeterminate in 5 cases (details Table 1 and Appendix 1).

### National reporting rate of hyper-inflammatory syndrome following mRNA vaccine

Considering all 9 cases of hyper-inflammatory syndrome, a national reporting rate of 1.1 (95% CI [0.5; 2.1]) per 1,000,000 mRNA vaccine doses administered in 12-17-year-old children was observed. Excluding cases for which evidence of previous SARS-CoV-2 infection has been found, the reporting rate was reduced to 0.7 (95% CI [0.3; 1.6]) per 1,000,000 mRNA vaccine doses administered. This reporting rate varied from 1.2 (95% CI [0.4; 2.9]) following the first mRNA vaccine injection to 1.0 (95% CI [0.3; 2.6]) after the second mRNA vaccine injection (Table 2). The reporting rate was significantly higher for males compared to females (1.9 [0.8; 3.8] versus 0.3 [0.0; 1.4] per 1,000,000 doses, respectively, p=0.039). As a comparator, 130 post-SARS-CoV-2 MIS-C cases occurred in 12-17-year-old children, for 1,148,299 same-age children infected by SARS-CoV-2, leading to a MIS-C rate of 113.3 [94.7; 134.6] per 1,000,000 12-17-year-old infected children (Table 2).

**Table 2:** Rate of hyper-inflammatory syndrome following COVID-19 mRNA vaccine compared to MIS-C post SARS-CoV-2 infection in 12-17-year-old children in France.

A) Reporting rate of hyper-inflammatory syndrome following COVID-19 mRNA vaccine in 12-17-year-old children
|  | Number of injected doses | Number of hyper-inflammatory syndrome | Reporting rate per 1,000,000 doses |
| --- | --- | --- | --- |
| Overall vaccination | 8,113,058 | 9 | 1.1 [0.5; 2.1] |
| Excluding cases for which evidence of previous SARS-CoV-2 infection has been found | 8,113,058 | 6 | 0.7 [0.3; 1.6] |
| First COVID-19 mRNA injection | 4,079,234 | 5 | 1.2 [0.4; 2.9] |
| Second COVID-19 mRNA injection | 3,905,636 | 4 | 1.0 [0.3; 2.6] |
| *Vaccination by sex* |  |  |  |
| Males | 4,126,275 | 8 | 1.9 [0.8; 3.8] |
| Females | 3,986,783 | 1 | 0.3 [0.0; 1.4] |

B) Rate of MIS-C following SARS-CoV-2 infection in 12-17-year-old children
|  | Estimated number of infected children | Number of MIS-C | Rate per 1,000,000 of infections |
| --- | --- | --- | --- |
|  | 1,147,150 | 130 | 113.3 [94.7; 134.6] |
| *MIS-C by sex* |  |  |  |
| Males | 587,086 | 74 | 126.0 [99.0; 158.2] |
| Females | 560,064 | 56 | 100.0 [75.5; 129.8] |

Abbreviations: MIS-C: multisystem inflammatory syndrome in children.

### Clinical features of hyper-inflammatory syndrome following COVID-19 mRNA vaccine compared to post-SARS-CoV-2 MIS-C cases

The detailed clinical presentation of children with hyper-inflammatory syndrome following COVID-19 mRNA vaccine is provided Table 1. Median age was 12.5 years (IQR [12.0; 13.5]), 8/9 (89%) children were male and 3/8 had comorbidities (one type 1 diabetes, one osteochondritis with overweight, and one leukemia in remission). The most frequent clinical features were cardiac involvement (8/9, 89%, including 7 elevated cardiac enzymes, 4 pericarditis, 3 acute left ventricular ejection fraction decrease ≤ 55%, 1 transient coronary dilation and 1 myocarditis), gastrointestinal symptoms (7/9, 78%), coagulopathy (5/9, 56%), mucocutaneous involvement (5/9, 56%), cytolytic hepatitis (4/9, 44%) and shock (3/9, 33%). Macrophage activation syndrome was identified in one case. For comparison, among 199 children with post-SARS-CoV-2 MIS-C, 108 (54%) were male (p=0.081), and cardiac involvement was found in 63% (p=0.16, Table 3).

**Table 3:** Comparison of clinic-biological features of hyper-inflammatory syndrome following COVID-19 mRNA vaccine and MIS-C post SARS-CoV-2 infection in France.

|  | Hyper-inflammatory syndrome following COVID-19 mRNA vaccine (N=9) | MIS-C post SARS-CoV-2 infection (N=199) | P value |
| --- | --- | --- | --- |
| **Clinical characteristics** |  |  |  |
| Sex ratio (F/M) | 0.13 | 0.84 | 0.081 |
| Age | 12.5 [12.0; 13.5] | 8.7 [4.8; 12.1] |  |
| *Organ involvement following MIS-C WHO definition* |  |  |  |
| Mucocutaneous involvement | 5 (56%) | 159 (80%) | 0.097 |
| Shock | 3 (33%) | 100 (50%) | 0.318 |
| Cardiac involvement | 8 (89%) | 125 (63%) | 0.161 |
| Including LVEF ≤ 55% | 3 (33%) | 52 (26%) | 0.701 |
| Coagulopathy | 5 (56%) | 82 (41%) | 0.496 |
| Digestive symptoms | 7 (78%) | 178 (89%) | 0.261 |
| *Other organ involvement* |  |  |  |
| Cytolytic hepatitis | 4 (44%) | 36 (18%) | 0.071 |
| Lymphadenopathy | 3 (33%) | 39 (20%) | 0.390 |
| Renal failure | 2 (22%) | 31 (16%) | 0.637 |
| Neurological involvement | 1 (13%) | 65 (33%) | 0.278 |
| **Biological features** |  |  |  |
| Maximal CRP, mg/L | 186.0 [97.0; 250.0] | 271.0 [187.2; 353.5] | **0.031** |
| Ferritinemia, (μg/L) | 527.0 [369.5; 4892.5] | 390.0 [206.0; 748.5] | 0.228 |
| Hemoglobin, g/dL | 12.3 [11.6; 13.6] | 11.0 [10.1; 11.0] | **0.022** |
| Leucocytes, /mm<sup>3</sup> | 10 560 [7 530; 13 720] | 9 945 [7 000; 14.155] | 0.768 |
| Neutrophils, /mm<sup>3</sup> | 9 400 [5 970; 11 350] | 8 090 [5 245; 12 065] | 0.940 |
| Lymphocytes, /mm<sup>3</sup> | 890 [530; 910] | 1 000 [690; 1 805] | 0.217 |
| Eosinophils, /mm<sup>3</sup> | 220 [15; 740] | 100 [5; 220] | 0.463 |
| Platelets, /mm<sup>3</sup> | 230 000 [204 000; 321 000] | 186 000 [142 000; 277 500] | 0.336 |
| **Short term outcomes** |  |  |  |
| PICU transfer | 3 (33%) | 143 (72%) | **0.022** |
| Hemodynamic support | 2 (22%) | 86 (43%) | 0.307 |
| Hospital length of stay | 7.0 [7.0; 9.0] | 8.0 [6.0; 12.0] | 0.960 |

Categorical variables are described with numbers (percentages) and quantitative variables are described with median (IQR). Abbreviations: MIS-C: multisystem inflammatory syndrome in children.

Some biological parameters differed between post-vaccine hyper-inflammatory syndrome and post-SARS-CoV-2 MIS-C, including inflammatory parameters (median CRP level 186 [97; 250] vs 271 [187; 354], respectively, p=0.031), and blood cell count (median hemoglobin 12.3 [11.6; 13.6] vs 11.0 [10.1; 11.0], respectively, p=0.022, Table 3). Of note, 2/9 (22%) children with post-vaccine hyper-inflammatory syndrome had a transient hypereosinophilia (compared to 9/199 (5%) in post-SARS-CoV-2 MIS-C). These two children were explored for TRBV11-2/Vb21.3 expansion, none of them had an expansion of this repertory, while it was present in 75% of post-SARS-CoV-2 MIS-C patients^11^.

### Specific therapy and short-term outcomes

Short term outcomes seemed also less severe for post-vaccine hyper-inflammatory syndrome, with a lower rate of PICU transfer (3/9, 33%), compared to 143/199 (72%) for post-SARS-CoV-2 MIS-C (p=0.022, Table 3). Six children were treated by an association of intravenous immunoglobulins plus methylprednisolone, of whom one received a subsequent 10mg/kg/day methylprednisolone pulse. Two children received methylprednisolone alone, and one did not receive specific immunomodulator agent. All children fully recovered at the time of discharge. Median length of hospital stay was 7 days (IQR [7; 9]).

## Discussion

To our knowledge, this is the first post-authorization population-based pharmacovigilance study assessing the risk of hyper-inflammatory syndrome following COVID-19 mRNA vaccine in 12-17-year-old children. We found that this entity was observed with an reporting rate of 1.1 (95% CI [0.5; 2.1]) per 1,000,000 doses in this population. In most cases, no evidence of previous SARS-CoV-2 infection was observed, suggesting a link between this entity and COVID-19 mRNA vaccine. This rare serious adverse event should be put in balance with the rate of post-SARS-CoV-2 MIS-C in the same age group in the same population, which was 100-fold higher. A recent study highlighted that COVID-19 mRNA vaccine may significantly reduce the incidence of post-SARS-CoV-2 MIS-C (Hazard Ratio 0.09 (95% CI, 0.04-0.21)).^31^ Taken together, these findings suggest that the benefit-risk balance of COVID-19 mRNA vaccine is largely in favor of the vaccination in this age group, in a context of active circulation of SARS-CoV-2.

An important issue is to delineate the clinical spectrum of this entity, which may have overlap with several other diseases. First, cases of myocarditis have been reported following COVID-19 mRNA vaccines, especially in young men.^36,37^ These cases mainly occurred after the second dose of vaccine, few days after the injection, and were rapidly resolutive in most cases.^36,37^ The higher rate of hyper-inflammatory syndrome following COVID-19 mRNA vaccine in males, and the rate of cardiac involvement (8/9 cases) suggest similitudes with this entity. However, myocarditis were classically afebrile, with low inflammatory parameters, and were a mono-organ involvement.^37^ These major clinical differences may allow distinguishing these two entities. Second, all cases of hyper-inflammatory syndrome following COVID-19 mRNA vaccine fulfilled WHO definition for MIS-C. Indeed, the prolonged hyper-inflammatory state, the multi-organ involvement and the severity of the disease are principal features of these two entities,^7^ indicating at least a major overlap. However, if statistical comparison between these two diseases was limited by the low number of cases, our findings suggest that post-SARS-CoV-2 MIS-C may have still higher inflammatory parameters, and more cytopenia. This may be in line with the significantly lower rate of PICU transfer (33% vs 72%) for hyper-inflammatory syndrome following COVID-19 mRNA vaccine cases, which might reflect a less severe immune storm and disease course. Notably, a 4-week delay has been observed in the context of MIS-C following SARS-CoV-2 infection^38–40^. Here, the delay from first antigen exposure to hyper-inflammatory syndrome occurred within a week in 3 patients and after 4-12 weeks in the 6 others. In the cases with early reaction, a hypereosinophilia was seen in 2 patients, a feature not seen in classical MIS-C. This observation might reflect immunoallergic reaction distinct from the superantigenic like features of MIS-C. Expansion of Vb21.3 expressing T cells is a hallmark of the MIS-C and can be easily assessed by flowcytometry.^11^ By contrast, the two children with post-vaccination hyper-inflammatory syndrome had no expansion of this T cell subset. Taken together, these clinical and immunological divergences may imply distinct underlying pathways and further studies are required to expand this finding. Third, a recent study coordinated by the CDC reported cases of multisystem inflammatory system in adults (MIS-A) in USA, in vaccinated and unvaccinated adults.^32^ Twenty cases were identified, of whom seven were vaccinated. However, all of them had a documented previous exposure to SARS-CoV-2, questioning the direct causal role of mRNA vaccines in these manifestations, and diverging with the pediatric syndrome reported here, with only 2/9 patients presenting a seropositivity to N antigen. The issue of delineating these different entities underline the need to extensively investigate cases of hyper-inflammatory syndromes following mRNA vaccines, especially by performing anti-S and anti-N serology, along with exploration for TRBV11-2/Vb21.3 expansion.

Because this hyper-inflammatory syndrome following COVID-19 mRNA vaccine was severe with acute multi-organ dysfunction, therapeutic aspects are of major interest. In this cohort, most children were treated by an association of immunoglobulins plus methylprednisolone, following MIS-C therapeutic protocols.^30^ Only one child treated with this combination required a therapeutic escalation, and received a methylprednisolone pulse (10mg/kg/day). All children fully recovered. If sample size precludes any definitive conclusion, these findings suggest that the association of immunoglobulins plus methylprednisolone may be a suitable approach while awaiting more evidence regarding these treatments.

Several limitations should be discussed. First the causality of COVID-19 mRNA vaccines assessment was mainly based on investigation for previous SARS-CoV-2 infection. However, pauci or asymptomatic infections are frequent in children, and may not have been documented. Furthermore, false negatives can be observed for anti-Nucleocapsid serology.^41^ Thus, we cannot exclude that some of the cases reported here could be related to undiagnosed SARS-CoV-2 infections. Second, because this entity has not been previously described in healthy populations, we could not have a control population to estimate the expected incidence of this disease in unvaccinated children, which would help in elucidating the vaccine causality.^16^ Third, we cannot rule out under reporting of adverse drug events in our population, which may have biased the estimated rate of hyper-inflammatory syndrome. However, following the implementation of COVID-19 mRNA vaccine, a major effort has been made by all pharmacovigilance centers to publicize that the reporting of any suspected adverse drug reaction following these vaccines was mandatory.^26^ The impressive number of suspected adverse drug reaction reports (>80,000 between January 2021 and January 2022 in France) suggest that underreporting may have been very rare, especially for serious adverse drug reactions.^26^ Fourth, given the very low proportion of 12-17-year-old children vaccinated by mRNA-1273 (<5%), we could not conduct subgroup analysis to compare the risk of hyper-inflammatory syndromes according to COVID-19 mRNA vaccine type. Further studies are required to explore if this risk differ between BNT162b2 and mRNA-1273. Fifth, because mRNA vaccines were only recommended to 12-17-year-old children in France until December 2021, we could not explore the risk of hyper-inflammatory syndrome in younger children.

## Conclusion

In this study, we identified cases of hyper-inflammatory syndrome following COVID-19 mRNA vaccines in 12-17-year-old children in France. This syndrome was very rare, and its reporting rate, in comparison with the rate of MIS-C following SARS-CoV-2 infection in the same age-group, largely supports the vaccination in a context of an important circulation of SARS-CoV-2. This syndrome shared many clinical features with post-SARS-CoV-2 MIS-C, but some clinical, immunological and short-term outcomes divergences call for further studies to explore its specific pathway.

## Data Availability

All data produced in the present study are available upon reasonable request to the authors

## Author Contributions

NO, HB, FA, and AB made substantial contributions to the conception or design of the work. NO and AB drafted the manuscript. NO, HB, CLeb, CLev, EJ, BBM, CO, SR, VH, IKP, ND, LDP, XI, CGLG, FA and AB were involved in the acquisition, analysis, or interpretation of data. All authors provided critical revision of the manuscript for important intellectual content. NO and HB had full access to all data in the study and take responsibility for the integrity of the data and the accuracy of the data analysis.

## Funding/Support

NO was supported by the 2021 ESPID (European Society for Pediatric Infectious Diseases) Fellowship Award. the French Covid-19 Paediatric Inflammation Consortium received an unrestricted grant from the Square Foundation (Grandir–Fonds de Solidarité Pour L’enfance).

## Conflicts of Interest Disclosures

NO reports travel grants from GSK, Pfizer, and Sanofi. Dr Javouhey reported receiving grants from CSL Behring. Dr C. Levy reported receiving grants from GlaxoSmithKline, Merck Sharp & Dohme, and Sanofi and personal fees from Pfizer and Merck. Dr Cohen reported receiving personal fees from GlaxoSmithKline, Pfizer, Sanofi, and Merck Sharp & Dohme. All other authors have no potential conflicts of interest to disclose.

## Role of the Funder/Sponsor

The funders had no role in the design or conduct of the study, collection management, analysis, or interpretation of the data; preparation, review, or approval of the manuscript; or the decision to submit the manuscript for publication.

## Additional Contributions

We are grateful to Agence Nationale de Sécurité du Médicament et des produits de santé (ANSM), Santé Publique France, Société Française de Pédiatrie, Groupe de Pédiatrie Générale, Groupe de Pathologie Infectieuse Pédiatrique, Groupe Francophone de Réanimation et d’Urgences Pédiatriques, Société Française de Cardiologie, Filiale de Cardiologie Pédiatrique et Congénitale, Société Francophone Dédiée à L’étude des Maladies Inflammatoires Pédiatriques, and Filière de Santé des Maladies Auto-immunes et Auto-inflammatoires Rares for their participation in the French Covid-19 Paediatric Inflammation Consortium study. We thank Youssef Shaim, Laurence Baril, Mehdi Benkebil, Samuel Crommelynck, Baptiste Jacquot from ANSM, Isabelle Ramay, BSc, Claire Prieur, BSc, Marine Borg, Aurore Prieur, BSc, Laura Meyet, LLM, Jéremy Levy, BSc, Stéphane Bechet, MSc, and Sofia Abbou, LLM, from ACTIV (Association Clinique et Thérapeutique Infantile du Val-de-Marne), Créteil, France; Cecile Hoffart, MSc, and Maxime Brussieux, BSc, from Clinical Research Centre, Centre Hospitalier Intercommunal de Créteil, Créteil, France; Daniel Levy-Bruhl, MD, Mireille Allemand, Scarlett Georges, BSc, Valerie Olie, PhD, Nolween Regnault, PhD, and Jerome Naud, PharmD, from Santé Publique France, Agence Nationale de Santé Publique, Saint-Maurice; Murielle Herasse, PhD, from Filière de Santé Des Maladies Auto-immunes et Auto-inflammatoires Rares (FAI2R), Lyon, France; and David Skurnik, PhD, from INSERM U1151-Equipe 11. We are grateful to every microbiological laboratory staff member who performed severe acute respiratory syndrome coronavirus 2 reverse transcriptase– polymerase chain reaction and antibody testing in each center. None of the persons listed here received compensation for their role in the study.

### In addition to the authors, the following collaborators participated to the “French Covid-19 Pediatric Inflammation Consortium”

Maelle Selegny (Amiens); Lucas Jeusset, Aurelie Donzeau, Sophie Lety, Bertrand Leboucher (Angers); Agnes Baur (Annecy); Cristian Fedorczuk (Arcachon); Marion Lajus, Philippe Bensaid (Argenteuil); Yacine Laoudi (Aulnay Sous Bois); Charlotte Pons (Avignon); Anne-Cécile Robert, Camille Beaucourt (Besançon); Loïc De Pontual (Bondy); Muriel Richard, Etienne Goisque, Xavier Iriart, Olivier Brissaud, Pierre Segretin, Julie Molimard (Bordeaux); Marie-Clothilde Orecel, Gregoire Benoit (Boulognes Billancourt); Lucille Bongiovanni (Brest); Guerder Margaux, Robin Pouyau, Jean-Marie De Guillebon De Resnes, Ellia Mezgueldi, Fleur Cour-Andlauer, Come Horvat, Pierre Poinsot, Cecile Frachette, Antoine Ouziel, Yves Gillet (Bron); Catherine Barrey (Bry Sur Marne); Jacques Brouard, Florence Villedieu (Caen); Vathanaksambath Ro, Narcisse Elanga (Cayenne); Vincent Gajdos (Clamart); Romain Basmaci (Colombes); Hadile Mutar (Contamine sur Arve); Sébastien Rouget (Corbeil Essone); Elodie Nattes, Isabelle Hau, Sandra Biscardi, El Jurdi Houmam, Camille Jung (Créteil); Denis Semama, Frederic Huet (Dijon); Anne-Marie Zoccarato (Gap); Mayssa Sarakbi (Gonnesse); Guillaume Mortamet, Cécile Bost-Bru (Grenoble); Joachim Bassil (Laval); Caroline Vinit, Véronique Hentgen (Le Chesnay); Pascal Leroux, Valérie Bertrand, Caroline Parrod (Le Havre); Irina Craiu, Isabelle Kone-Paut, Philippe Durand, Pierre Tissiere, Caroline Claude, Guillaume Morelle, Tamazoust Guiddir, Charlotte Borocco (Le Kremlin-Bicêtre); Frédérique Delion (Les Abymes); Camille Guillot, Stéphane Leteurtre, François Dubos, Mylene Jouancastay, Alain Martinot, Valentine Voeusler (Lilles); Jane Languepin (Limoges); Nathalie Garrec, Arnaud Chalvon Demersay (Marne La Vallée); Aurélie Morand, Emmanuelle Bosdure, Noémie Vanel, Fabrice Ughetto, Fabrice Michel (Marseille); Caujolle Marie, Renaud Blonde, Jacqueline Nguyen (Mayotte); Olivier Vignaud, Caroline Masserot-Lureau, François Gouraud, Carine Araujo (Meaux); Tara Ingrao (Metz); Sanaa Naji (Mont de Marsans); Mohammed Sehaba (Montargis); Christine Roche (Montbrison); Aurelia Carbasse, Christophe Milesi (Montpellier); Mustapha Mazeghrane (Montreuil); Sandrine Haupt (Mulhouse); Cyril Schweitzer (Nancy); Benedicte Romefort, Elise Launay, Christèle Gras-Le Guen (Nantes); Ahmed Ali, Nathalie Blot (Neuilly Sur Seine); Antoine Tran, Anne Rancurel, Mickael Afanetti (Nice); Sophie Odorico (Nîmes); Deborah Talmud (Orléans); Anais Chosidow, Anne-Sophie Romain, Emmanuel Grimprel Marie Pouletty, Jean Gaschignard, Olivier Corseri, Albert Faye, Jean Gaschignard, Isabelle Melki, Camille Ducrocq, Cherine Benzoïd, Johanna Lokmer, Stéphane Dauger, Maryline Chomton, Anna Deho, Fleur Lebourgeois, Sylvain Renolleau, Fabrice Lesage, Florence Moulin, Laurent Dupic, Yael Pinhas, Agathe Debray, Martin Chalumeau, Véronique Abadie, Pierre Frange, Jeremie F Cohen, Slimane Allali, William Curtis, Zahra Belhadjer, Johanne Auriau, Mathilde Méot, Lucile Houyel, Damien Bonnet, Christophe Delacourt, Brigitte Bader Meunier, Pierre Quartier, Youssef Shaim, Laurence Baril, Samuel Crommelynck, Baptiste Jacquot (Paris); Philippe Blanc (Poissy); Natacha Maledon (Poitiers); Blandine Robert (Pontoise); Camille Loeile (Quimper); Clémence Cazau, Gauthier Loron (Reims); Simona Gaga (Remiremont); Cécile Vittot, Loubna El Nabhani (Rouen); François Buisson (Saumur); Muriel Prudent (Sens); Hugues Flodrops (St Denis, La Réunion); Fadhila Mokraoui, Simon Escoda (St Denis); Nina Deschamps (St Malot); Laurent Bonnemains, Sarah-Louisa Mahi, Clara Mertes, Joelle Terzic, Julie Helms (Strasbourg); Charlotte Idier (Tours); Soraya Chenichene, Nicoleta Magdolena Ursulescu (Trévenans); Gladys Beaujour (Villeneuve Saint Georges).

### In addition to the authors, the following collaborators participated to the “French Pharmacovigilance network”

Anaïs Gaiffe (Besançon), Antoine Pariente, Francesco Salvo (Bordeaux), Joelle Micallef (Marseille), Martine Tebacher (Strasbourg), Haleh Bagheri (Toulouse), Marie-Sarah Agier (Tours).

## Notes

### Author Declarations

The study was approved by the French INSERM ethics committee for evaluation (IRB00003888). A written information form validated by the ethics committee was given to all participants. Oral consent was obtained from study participants; no family members or participants refused to participate.

