## Supplemental material for "Multisystemic inflammatory syndrome following COVID-19 mRNA vaccine in children: a national post-authorization pharmacovigilance study"

Appendix 1: Worksheet for adverse event following immunization causality assessment following WHO 2019 guidelines^1^

Step 1 (Eligibility)

| *Patient ID/Name :* ***Case 1.*** |  | *DoB/Age: 12 years.* |  | *Sex: Male* |
| --- | --- | --- | --- | --- |
| Name one of the vaccines administered before this event |  | What is the Valid Diagnosis? |  | Does the diagnosis meet a case definition? |
| BNT162b2 |  | Hyper-inflammatory syndrome |  | Yes (WHO MIS-C definition) |
| **Create your question on causality here**  Has the BNT162b2 vaccine / vaccination caused Hyper-inflammatory syndrome (The event for review  in step 2 - valid diagnosis) | | | | |
| Is this case eligible for causality assessment? Yes; If, “Yes”, proceed to step 2 | | | | |

Step 2 (Event Checklist) ✓ (check) all boxes that apply

|  | Y | N | UK | NA | Remarks |
| --- | --- | --- | --- | --- | --- |
| I. Is there strong evidence for other causes? | | | | | |
| 1. In this patient, does the medical history, clinical examination and/or investigations, confirm another cause for the event? | 🞎 | ⌧ | 🞎 | 🞎 | No evidence of microbial cause of inflammation |
| II. Is there a known causal association with the vaccine or vaccination? | | | | | |
| *Vaccine product* | | | | | |
| 1. Is there evidence in published peer reviewed literature that this vaccine may cause such an event if administered correctly? | ⌧ | 🞎 | 🞎 | 🞎 | Several case reports published^2–5^ |
| 2. Is there a biological plausibility that this vaccine could cause such an event? | ⌧ | 🞎 | 🞎 | 🞎 | Exposure to Spike protein antigens |
| 3. In this patient, did a specific test demonstrate the causal role of the vaccine ? | 🞎 | 🞎 | 🞎 | ⌧ | No specific test for this syndrome |
| *Vaccine quality* | | | | | |
| 4. Could the vaccine given to this patient have a quality defect or is substandard or falsified? | 🞎 | ⌧ | 🞎 | 🞎 |  |
| *Immunization error* | | | | | |
| 5. In this patient, was there an error in prescribing or non-adherence to recommendations for use of the vaccine (e.g. use beyond the expiry date, wrong recipient etc.)? | 🞎 | ⌧ | 🞎 | 🞎 |  |
| 6. In this patient, was the vaccine (or diluent) administered in an unsterile manner? | 🞎 | ⌧ | 🞎 | 🞎 |  |
| 7. In this patient, was the vaccine’s physical condition (e.g. colour, turbidity, presence of foreign substances etc.) abnormal when administered? | 🞎 | ⌧ | 🞎 | 🞎 |  |
| 8. When this patient was vaccinated, was there an error in vaccine constitution/ preparation by the vaccinator (e.g. wrong product, wrong diluent, improper mixing, improper syringe filling etc.)? | 🞎 | ⌧ | 🞎 | 🞎 |  |
| 9. In this patient, was there an error in vaccine handling (e.g. a break in the cold chain during transport, storage and/or immunization session etc.)? | 🞎 | ⌧ | 🞎 | 🞎 |  |
| 10. In this patient, was the vaccine administered incorrectly (e.g. wrong dose, site or route of administration; wrong needle size etc.)? | 🞎 | ⌧ | 🞎 | 🞎 |  |
| *Immunization anxiety (Immunization stress related responses - ISRR)* | | | | | |
| 11. In this patient, could this event be a stress response triggered by immunization (e.g. acute stress response, vasovagal reaction, hyperventilation, dissociative neurological symptom reaction etc)? | 🞎 | ⌧ | 🞎 | 🞎 |  |
| II (time): Was the event in section II within the time window of increased risk (i.e. ‘Yes” response to questions from II 1 to II 11 above) | | | | | |
| 12. In this patient, did the event occur within a plausible time window after vaccine administration? | ⌧ | 🞎 | 🞎 | 🞎 |  |
| III. Is there strong evidence against a causal association? | | | | | |
| 1. Is there a body of published evidence (systematic reviews, GACVS reviews, Cochrane reviews etc.) **against** a causal association between the vaccine and the event? | 🞎 | ⌧ | 🞎 | 🞎 | No population based or pharmacovigilance study infirming the risk of hyper-inflammatory syndrome following mRNA COVID-19 vaccines in children |
| IV. Other qualifying factors for classification | | | | | |
| 1. In this patient, did such an event occur in the past after administration of a similar vaccine? | 🞎 | ⌧ | 🞎 | 🞎 |  |
| 2. In this patient, did such an event occur in the past independent of vaccination? | 🞎 | ⌧ | 🞎 | 🞎 |  |
| 3. Could the current event have occurred in this patient without vaccination (background rate)? | 🞎 | ⌧ | 🞎 | 🞎 |  |
| 4. Did this patient have an illness, pre-existing condition or risk factor that could have contributed to the event? | 🞎 | ⌧ | 🞎 | 🞎 |  |
| 5. Was this patient taking any medication prior to the vaccination? | 🞎 | ⌧ | 🞎 | 🞎 |  |
| 6. Was this patient exposed to a potential factor (other than vaccine) prior to the event (e.g. allergen, drug, herbal product etc.)? | 🞎 | ⌧ | 🞎 | 🞎 | Especially, no evidence of previous exposure to SARS-CoV-2 |

Note: Y: Yes; N: No; UK: Unknown; NA: Not applicable.

No

Step 3 (Algorithm) review all steps and ✓ all the appropriate boxes

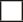

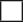

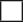

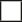

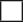

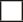

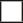

I A. Inconsistent causal association to immunization

III A. Inconsistent causal association to immunization

Yes

Yes

I. Is there strong evidence for other

causes?

II. Is there a known causal association with the vaccine/

vaccination

III. Is there a strong evidence against a causal association?

IV. Review other qualifying factors

Yes

II (Time). Was the event within the time window of increased risk?

Is the event classifiable?

IV D. Unclassifiable

Yes

Yes

II A. Consistent causal association to immunization

IV A. Consistent causal association to immunization

IV B. Indeterminate

IV C. Inconsistent causal association to immunization

Mandatory path

**Notes for Step 3:**

II A: With the available information, it seems likely that the vaccine could be involved in the event (no evidence of other cause, similar cases described in the literature, time window plausible)

IV A: No strong evidence against a causal association, and no previous exposure to a potential factor, i.e., no evidence of SARS-CoV-2 infection despite extensive investigation.

Step 4 (Classification) ✓ all boxes that apply

Adequate information available

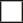

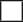

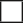

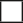

A. Consistent with causal association to immunization

A4. Immunization anxiety-related reaction (ISRR**)

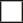

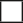

B. Indeterminate

B2. Qualifying factors result in conflicting trends of consistency and inconsistency with causal association to immunization

B1. *Temporal relationship is consistent but there is insuficient definitive evidence for vaccine causing event (may be new vaccine-linked event)

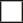

C. Inconsistent with causal association to immunization

C. Coincidental Underlying or emerging

condition(s), or condition(s) caused by exposure to something other than vaccine

A1. Vaccine product-related reaction

(As per published literature)

A2. Vaccine quality defect-related reaction

A3. Immunization error-related reaction

Adequate information not available

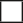

Unclassifiable

Specify the additional information required for classification :

**B1 : Potential signal and maybe considered for investigation*

*** Immunization stress related response*

**Summarize the classification logic in the order of priority:**

With available evidence, we could conclude that the classification is consistent because: there is no evidence of an other cause for the event (especially, no evidence of previous SARS-CoV-2 infection despite extensive investigation), similar cases have been described in the literature, the time window is compatible, there is no strong evidence against a causal association.

Step 1 (Eligibility)

| *Patient ID/Name :* ***Case 2.*** |  | *DoB/Age: 15 years.* |  | *Sex: Male* |
| --- | --- | --- | --- | --- |
| Name one of the vaccines administered before this event |  | What is the Valid Diagnosis? |  | Does the diagnosis meet a case definition? |
| BNT162b2 |  | Hyper-inflammatory syndrome |  | Yes (WHO MIS-C definition) |
| **Create your question on causality here**  Has the BNT162b2 vaccine / vaccination caused Hyper-inflammatory syndrome (The event for review  in step 2 - valid diagnosis) | | | | |
| Is this case eligible for causality assessment? Yes; If, “Yes”, proceed to step 2 | | | | |

Step 2 (Event Checklist) ✓ (check) all boxes that apply

|  | Y | N | UK | NA | Remarks |
| --- | --- | --- | --- | --- | --- |
| I. Is there strong evidence for other causes? | | | | | |
| 1. In this patient, does the medical history, clinical examination and/or investigations, confirm another cause for the event? | 🞎 | ⌧ | 🞎 | 🞎 | No evidence of microbial cause of inflammation |
| II. Is there a known causal association with the vaccine or vaccination? | | | | | |
| *Vaccine product* | | | | | |
| 1. Is there evidence in published peer reviewed literature that this vaccine may cause such an event if administered correctly? | ⌧ | 🞎 | 🞎 | 🞎 | Several case reports published^2–5^ |
| 2. Is there a biological plausibility that this vaccine could cause such an event? | ⌧ | 🞎 | 🞎 | 🞎 | Exposure to Spike protein antigens |
| 3. In this patient, did a specific test demonstrate the causal role of the vaccine ? | 🞎 | 🞎 | 🞎 | ⌧ | No specific test for this syndrome |
| *Vaccine quality* | | | | | |
| 4. Could the vaccine given to this patient have a quality defect or is substandard or falsified? | 🞎 | ⌧ | 🞎 | 🞎 |  |
| *Immunization error* | | | | | |
| 5. In this patient, was there an error in prescribing or non-adherence to recommendations for use of the vaccine (e.g. use beyond the expiry date, wrong recipient etc.)? | 🞎 | ⌧ | 🞎 | 🞎 |  |
| 6. In this patient, was the vaccine (or diluent) administered in an unsterile manner? | 🞎 | ⌧ | 🞎 | 🞎 |  |
| 7. In this patient, was the vaccine’s physical condition (e.g. colour, turbidity, presence of foreign substances etc.) abnormal when administered? | 🞎 | ⌧ | 🞎 | 🞎 |  |
| 8. When this patient was vaccinated, was there an error in vaccine constitution/ preparation by the vaccinator (e.g. wrong product, wrong diluent, improper mixing, improper syringe filling etc.)? | 🞎 | ⌧ | 🞎 | 🞎 |  |
| 9. In this patient, was there an error in vaccine handling (e.g. a break in the cold chain during transport, storage and/or immunization session etc.)? | 🞎 | ⌧ | 🞎 | 🞎 |  |
| 10. In this patient, was the vaccine administered incorrectly (e.g. wrong dose, site or route of administration; wrong needle size etc.)? | 🞎 | ⌧ | 🞎 | 🞎 |  |
| *Immunization anxiety (Immunization stress related responses - ISRR)* | | | | | |
| 11. In this patient, could this event be a stress response triggered by immunization (e.g. acute stress response, vasovagal reaction, hyperventilation, dissociative neurological symptom reaction etc)? | 🞎 | ⌧ | 🞎 | 🞎 |  |
| II (time): Was the event in section II within the time window of increased risk (i.e. ‘Yes” response to questions from II 1 to II 11 above) | | | | | |
| 12. In this patient, did the event occur within a plausible time window after vaccine administration? | ⌧ | 🞎 | 🞎 | 🞎 |  |
| III. Is there strong evidence against a causal association? | | | | | |
| 1. Is there a body of published evidence (systematic reviews, GACVS reviews, Cochrane reviews etc.) **against** a causal association between the vaccine and the event? | 🞎 | ⌧ | 🞎 | 🞎 | No population based or pharmacovigilance study infirming the risk of hyper-inflammatory syndrome following mRNA COVID-19 vaccines in children |
| IV. Other qualifying factors for classification | | | | | |
| 1. In this patient, did such an event occur in the past after administration of a similar vaccine? | 🞎 | ⌧ | 🞎 | 🞎 |  |
| 2. In this patient, did such an event occur in the past independent of vaccination? | 🞎 | ⌧ | 🞎 | 🞎 |  |
| 3. Could the current event have occurred in this patient without vaccination (background rate)? | 🞎 | ⌧ | 🞎 | 🞎 |  |
| 4. Did this patient have an illness, pre-existing condition or risk factor that could have contributed to the event? | 🞎 | ⌧ | 🞎 | 🞎 |  |
| 5. Was this patient taking any medication prior to the vaccination? | 🞎 | ⌧ | 🞎 | 🞎 |  |
| 6. Was this patient exposed to a potential factor (other than vaccine) prior to the event (e.g. allergen, drug, herbal product etc.)? | 🞎 | ⌧ | 🞎 | 🞎 | Especially, no evidence of previous exposure to SARS-CoV-2 |

Note: Y: Yes; N: No; UK: Unknown; NA: Not applicable.

No

Step 3 (Algorithm) review all steps and ✓ all the appropriate boxes

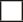

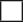

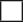

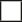

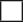

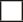

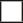

I A. Inconsistent causal association to immunization

III A. Inconsistent causal association to immunization

Yes

Yes

I. Is there strong evidence for other

causes?

II. Is there a known causal association with the vaccine/

vaccination

III. Is there a strong evidence against a causal association?

IV. Review other qualifying factors

Yes

II (Time). Was the event within the time window of increased risk?

Is the event classifiable?

IV D. Unclassifiable

Yes

Yes

II A. Consistent causal association to immunization

IV A. Consistent causal association to immunization

IV B. Indeterminate

IV C. Inconsistent causal association to immunization

Mandatory path

**Notes for Step 3:**

II A: With the available information, it seems likely that the vaccine could be involved in the event (no evidence of other cause, similar cases described in the literature, time window plausible)

IV A: No strong evidence against a causal association, and no previous exposure to a potential factor, i.e., no evidence of SARS-CoV-2 infection despite extensive investigation.

Step 4 (Classification) ✓ all boxes that apply

Adequate information available

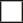

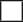

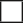

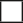

A. Consistent with causal association to immunization

A4. Immunization anxiety-related reaction (ISRR**)

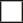

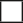

B. Indeterminate

B2. Qualifying factors result in conflicting trends of consistency and inconsistency with causal association to immunization

B1. *Temporal relationship is consistent but there is insuficient definitive evidence for vaccine causing event (may be new vaccine-linked event)

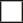

C. Inconsistent with causal association to immunization

C. Coincidental Underlying or emerging

condition(s), or condition(s) caused by exposure to something other than vaccine

A1. Vaccine product-related reaction

(As per published literature)

A2. Vaccine quality defect-related reaction

A3. Immunization error-related reaction

Adequate information not available

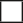

Unclassifiable

Specify the additional information required for classification :

**B1 : Potential signal and maybe considered for investigation*

*** Immunization stress related response*

**Summarize the classification logic in the order of priority:**

With available evidence, we could conclude that the classification is consistent because: there is no evidence of an other cause for the event (especially, no evidence of previous SARS-CoV-2 infection despite extensive investigation), similar cases have been described in the literature, the time window is compatible, there is no strong evidence against a causal association.

Step 1 (Eligibility)

| *Patient ID/Name :* ***Case 3.*** |  | *DoB/Age: 16 years.* |  | *Sex: Male* |
| --- | --- | --- | --- | --- |
| Name one of the vaccines administered before this event |  | What is the Valid Diagnosis? |  | Does the diagnosis meet a case definition? |
| BNT162b2 |  | Hyper-inflammatory syndrome |  | Yes (WHO MIS-C definition) |
| **Create your question on causality here**  Has the BNT162b2 vaccine / vaccination caused Hyper-inflammatory syndrome (The event for review  in step 2 - valid diagnosis) | | | | |
| Is this case eligible for causality assessment? Yes; If, “Yes”, proceed to step 2 | | | | |

Step 2 (Event Checklist) ✓ (check) all boxes that apply

|  | Y | N | UK | NA | Remarks |
| --- | --- | --- | --- | --- | --- |
| I. Is there strong evidence for other causes? | | | | | |
| 1. In this patient, does the medical history, clinical examination and/or investigations, confirm another cause for the event? | 🞎 | ⌧ | 🞎 | 🞎 | No evidence of microbial cause of inflammation |
| II. Is there a known causal association with the vaccine or vaccination? | | | | | |
| *Vaccine product* | | | | | |
| 1. Is there evidence in published peer reviewed literature that this vaccine may cause such an event if administered correctly? | ⌧ | 🞎 | 🞎 | 🞎 | Several case reports published^2–5^ |
| 2. Is there a biological plausibility that this vaccine could cause such an event? | ⌧ | 🞎 | 🞎 | 🞎 | Exposure to Spike protein antigens |
| 3. In this patient, did a specific test demonstrate the causal role of the vaccine ? | 🞎 | 🞎 | 🞎 | ⌧ | No specific test for this syndrome |
| *Vaccine quality* | | | | | |
| 4. Could the vaccine given to this patient have a quality defect or is substandard or falsified? | 🞎 | ⌧ | 🞎 | 🞎 |  |
| *Immunization error* | | | | | |
| 5. In this patient, was there an error in prescribing or non-adherence to recommendations for use of the vaccine (e.g. use beyond the expiry date, wrong recipient etc.)? | 🞎 | ⌧ | 🞎 | 🞎 |  |
| 6. In this patient, was the vaccine (or diluent) administered in an unsterile manner? | 🞎 | ⌧ | 🞎 | 🞎 |  |
| 7. In this patient, was the vaccine’s physical condition (e.g. colour, turbidity, presence of foreign substances etc.) abnormal when administered? | 🞎 | ⌧ | 🞎 | 🞎 |  |
| 8. When this patient was vaccinated, was there an error in vaccine constitution/ preparation by the vaccinator (e.g. wrong product, wrong diluent, improper mixing, improper syringe filling etc.)? | 🞎 | ⌧ | 🞎 | 🞎 |  |
| 9. In this patient, was there an error in vaccine handling (e.g. a break in the cold chain during transport, storage and/or immunization session etc.)? | 🞎 | ⌧ | 🞎 | 🞎 |  |
| 10. In this patient, was the vaccine administered incorrectly (e.g. wrong dose, site or route of administration; wrong needle size etc.)? | 🞎 | ⌧ | 🞎 | 🞎 |  |
| *Immunization anxiety (Immunization stress related responses - ISRR)* | | | | | |
| 11. In this patient, could this event be a stress response triggered by immunization (e.g. acute stress response, vasovagal reaction, hyperventilation, dissociative neurological symptom reaction etc)? | 🞎 | ⌧ | 🞎 | 🞎 |  |
| II (time): Was the event in section II within the time window of increased risk (i.e. ‘Yes” response to questions from II 1 to II 11 above) | | | | | |
| 12. In this patient, did the event occur within a plausible time window after vaccine administration? | ⌧ | 🞎 | 🞎 | 🞎 |  |
| III. Is there strong evidence against a causal association? | | | | | |
| 1. Is there a body of published evidence (systematic reviews, GACVS reviews, Cochrane reviews etc.) **against** a causal association between the vaccine and the event? | 🞎 | ⌧ | 🞎 | 🞎 | No population based or pharmacovigilance study infirming the risk of hyper-inflammatory syndrome following mRNA COVID-19 vaccines in children |
| IV. Other qualifying factors for classification | | | | | |
| 1. In this patient, did such an event occur in the past after administration of a similar vaccine? | 🞎 | ⌧ | 🞎 | 🞎 |  |
| 2. In this patient, did such an event occur in the past independent of vaccination? | 🞎 | ⌧ | 🞎 | 🞎 |  |
| 3. Could the current event have occurred in this patient without vaccination (background rate)? | 🞎 | ⌧ | 🞎 | 🞎 |  |
| 4. Did this patient have an illness, pre-existing condition or risk factor that could have contributed to the event? | 🞎 | ⌧ | 🞎 | 🞎 |  |
| 5. Was this patient taking any medication prior to the vaccination? | 🞎 | ⌧ | 🞎 | 🞎 |  |
| 6. Was this patient exposed to a potential factor (other than vaccine) prior to the event (e.g. allergen, drug, herbal product etc.)? | ⌧ | 🞎 | 🞎 | 🞎 | Evidence of previous exposure to SARS-CoV-2. |

Note: Y: Yes; N: No; UK: Unknown; NA: Not applicable.

No

Step 3 (Algorithm) review all steps and ✓ all the appropriate boxes

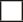

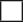

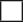

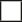

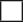

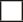

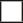

I A. Inconsistent causal association to immunization

III A. Inconsistent causal association to immunization

Yes

Yes

I. Is there strong evidence for other

causes?

II. Is there a known causal association with the vaccine/

vaccination

III. Is there a strong evidence against a causal association?

IV. Review other qualifying factors

Yes

II (Time). Was the event within the time window of increased risk?

Is the event classifiable?

IV D. Unclassifiable

Yes

Yes

II A. Consistent causal association to immunization

IV A. Consistent causal association to immunization

IV B. Indeterminate

IV C. Inconsistent causal association to immunization

Mandatory path

**Notes for Step 3:**

I A: With the available information, it seems that a potential other cause for the event has been identified (previous SARS-CoV-2 infection).

IV B: While there is a potential other cause for the event has been identified (previous SARS-CoV-2 infection), the potential combined role of SARS-CoV-2 exposure and mRNA COVID-19 vaccine exposure cannot be excluded.

Step 4 (Classification) ✓ all boxes that apply

Adequate information available

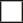

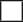

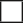

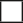

A. Consistent with causal association to immunization

A4. Immunization anxiety-related reaction (ISRR**)

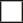

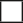

B. Indeterminate

B2. Qualifying factors result in conflicting trends of consistency and inconsistency with causal association to immunization

B1. *Temporal relationship is consistent but there is insuficient definitive evidence for vaccine causing event (may be new vaccine-linked event)

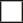

C. Inconsistent with causal association to immunization

C. Coincidental Underlying or emerging

condition(s), or condition(s) caused by exposure to something other than vaccine

A1. Vaccine product-related reaction

(As per published literature)

A2. Vaccine quality defect-related reaction

A3. Immunization error-related reaction

Adequate information not available

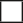

Unclassifiable

Specify the additional information required for classification :

**B1 : Potential signal and maybe considered for investigation*

*** Immunization stress related response*

**Summarize the classification logic in the order of priority:**

With available evidence, we could conclude that the classification is indeterminate because: there is evidence of an other potential cause for the event (evidence of previous SARS-CoV-2 infection), and the potential combined role of SARS-CoV-2 exposure and mRNA COVID-19 vaccine exposure cannot be excluded.

Step 1 (Eligibility)

| *Patient ID/Name :* ***Case 4.*** |  | *DoB/Age: 13 years.* |  | *Sex: Male* |
| --- | --- | --- | --- | --- |
| Name one of the vaccines administered before this event |  | What is the Valid Diagnosis? |  | Does the diagnosis meet a case definition? |
| BNT162b2 |  | Hyper-inflammatory syndrome |  | Yes (WHO MIS-C definition) |
| **Create your question on causality here**  Has the BNT162b2 vaccine / vaccination caused Hyper-inflammatory syndrome (The event for review  in step 2 - valid diagnosis) | | | | |
| Is this case eligible for causality assessment? Yes; If, “Yes”, proceed to step 2 | | | | |

Step 2 (Event Checklist) ✓ (check) all boxes that apply

|  | Y | N | UK | NA | Remarks |
| --- | --- | --- | --- | --- | --- |
| I. Is there strong evidence for other causes? | | | | | |
| 1. In this patient, does the medical history, clinical examination and/or investigations, confirm another cause for the event? | ⌧ | 🞎 | 🞎 | 🞎 | Evidence of previous SARS-CoV-2 infection |
| II. Is there a known causal association with the vaccine or vaccination? | | | | | |
| *Vaccine product* | | | | | |
| 1. Is there evidence in published peer reviewed literature that this vaccine may cause such an event if administered correctly? | ⌧ | 🞎 | 🞎 | 🞎 | Several case reports published^2–5^ |
| 2. Is there a biological plausibility that this vaccine could cause such an event? | ⌧ | 🞎 | 🞎 | 🞎 | Exposure to Spike protein antigens |
| 3. In this patient, did a specific test demonstrate the causal role of the vaccine ? | 🞎 | 🞎 | 🞎 | ⌧ | No specific test for this syndrome |
| *Vaccine quality* | | | | | |
| 4. Could the vaccine given to this patient have a quality defect or is substandard or falsified? | 🞎 | ⌧ | 🞎 | 🞎 |  |
| *Immunization error* | | | | | |
| 5. In this patient, was there an error in prescribing or non-adherence to recommendations for use of the vaccine (e.g. use beyond the expiry date, wrong recipient etc.)? | 🞎 | ⌧ | 🞎 | 🞎 |  |
| 6. In this patient, was the vaccine (or diluent) administered in an unsterile manner? | 🞎 | ⌧ | 🞎 | 🞎 |  |
| 7. In this patient, was the vaccine’s physical condition (e.g. colour, turbidity, presence of foreign substances etc.) abnormal when administered? | 🞎 | ⌧ | 🞎 | 🞎 |  |
| 8. When this patient was vaccinated, was there an error in vaccine constitution/ preparation by the vaccinator (e.g. wrong product, wrong diluent, improper mixing, improper syringe filling etc.)? | 🞎 | ⌧ | 🞎 | 🞎 |  |
| 9. In this patient, was there an error in vaccine handling (e.g. a break in the cold chain during transport, storage and/or immunization session etc.)? | 🞎 | ⌧ | 🞎 | 🞎 |  |
| 10. In this patient, was the vaccine administered incorrectly (e.g. wrong dose, site or route of administration; wrong needle size etc.)? | 🞎 | ⌧ | 🞎 | 🞎 |  |
| *Immunization anxiety (Immunization stress related responses - ISRR)* | | | | | |
| 11. In this patient, could this event be a stress response triggered by immunization (e.g. acute stress response, vasovagal reaction, hyperventilation, dissociative neurological symptom reaction etc)? | 🞎 | ⌧ | 🞎 | 🞎 |  |
| II (time): Was the event in section II within the time window of increased risk (i.e. ‘Yes” response to questions from II 1 to II 11 above) | | | | | |
| 12. In this patient, did the event occur within a plausible time window after vaccine administration? | ⌧ | 🞎 | 🞎 | 🞎 |  |
| III. Is there strong evidence against a causal association? | | | | | |
| 1. Is there a body of published evidence (systematic reviews, GACVS reviews, Cochrane reviews etc.) **against** a causal association between the vaccine and the event? | 🞎 | ⌧ | 🞎 | 🞎 | No population based or pharmacovigilance study infirming the risk of hyper-inflammatory syndrome following mRNA COVID-19 vaccines in children |
| IV. Other qualifying factors for classification | | | | | |
| 1. In this patient, did such an event occur in the past after administration of a similar vaccine? | 🞎 | ⌧ | 🞎 | 🞎 |  |
| 2. In this patient, did such an event occur in the past independent of vaccination? | 🞎 | ⌧ | 🞎 | 🞎 |  |
| 3. Could the current event have occurred in this patient without vaccination (background rate)? | 🞎 | ⌧ | 🞎 | 🞎 |  |
| 4. Did this patient have an illness, pre-existing condition or risk factor that could have contributed to the event? | 🞎 | ⌧ | 🞎 | 🞎 |  |
| 5. Was this patient taking any medication prior to the vaccination? | 🞎 | ⌧ | 🞎 | 🞎 |  |
| 6. Was this patient exposed to a potential factor (other than vaccine) prior to the event (e.g. allergen, drug, herbal product etc.)? | ⌧ | 🞎 | 🞎 | 🞎 | Evidence of previous exposure to SARS-CoV-2. |

Note: Y: Yes; N: No; UK: Unknown; NA: Not applicable.

No

Step 3 (Algorithm) review all steps and ✓ all the appropriate boxes

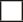

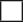

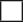

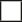

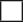

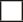

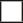

I A. Inconsistent causal association to immunization

III A. Inconsistent causal association to immunization

Yes

Yes

I. Is there strong evidence for other

causes?

II. Is there a known causal association with the vaccine/

vaccination

III. Is there a strong evidence against a causal association?

IV. Review other qualifying factors

Yes

II (Time). Was the event within the time window of increased risk?

Is the event classifiable?

IV D. Unclassifiable

Yes

Yes

II A. Consistent causal association to immunization

IV A. Consistent causal association to immunization

IV B. Indeterminate

IV C. Inconsistent causal association to immunization

Mandatory path

**Notes for Step 3:**

I A: With the available information, it seems that a potential other cause for the event has been identified (previous SARS-CoV-2 infection).

IV B: While there is a potential other cause for the event has been identified (previous SARS-CoV-2 infection), the potential combined role of SARS-CoV-2 exposure and mRNA COVID-19 vaccine exposure cannot be excluded.

Step 4 (Classification) ✓ all boxes that apply

Adequate information available

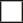

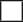

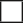

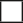

A. Consistent with causal association to immunization

A4. Immunization anxiety-related reaction (ISRR**)

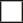

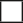

B. Indeterminate

B2. Qualifying factors result in conflicting trends of consistency and inconsistency with causal association to immunization

B1. *Temporal relationship is consistent but there is insuficient definitive evidence for vaccine causing event (may be new vaccine-linked event)

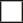

C. Inconsistent with causal association to immunization

C. Coincidental Underlying or emerging

condition(s), or condition(s) caused by exposure to something other than vaccine

A1. Vaccine product-related reaction

(As per published literature)

A2. Vaccine quality defect-related reaction

A3. Immunization error-related reaction

Adequate information not available

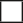

Unclassifiable

Specify the additional information required for classification :

**B1 : Potential signal and maybe considered for investigation*

*** Immunization stress related response*

**Summarize the classification logic in the order of priority:**

With available evidence, we could conclude that the classification is indeterminate because: there is evidence of an other potential cause for the event (evidence of previous SARS-CoV-2 infection), and the potential combined role of SARS-CoV-2 exposure and mRNA COVID-19 vaccine exposure cannot be excluded.

Step 1 (Eligibility)

| *Patient ID/Name :* ***Case 5.*** |  | *DoB/Age: 12 years.* |  | *Sex: Male* |
| --- | --- | --- | --- | --- |
| Name one of the vaccines administered before this event |  | What is the Valid Diagnosis? |  | Does the diagnosis meet a case definition? |
| BNT162b2 |  | Hyper-inflammatory syndrome |  | Yes (WHO MIS-C definition) |
| **Create your question on causality here**  Has the BNT162b2 vaccine / vaccination caused Hyper-inflammatory syndrome (The event for review  in step 2 - valid diagnosis) | | | | |
| Is this case eligible for causality assessment? Yes; If, “Yes”, proceed to step 2 | | | | |

Step 2 (Event Checklist) ✓ (check) all boxes that apply

|  | Y | N | UK | NA | Remarks |
| --- | --- | --- | --- | --- | --- |
| I. Is there strong evidence for other causes? | | | | | |
| 1. In this patient, does the medical history, clinical examination and/or investigations, confirm another cause for the event? | 🞎 | ⌧ | 🞎 | 🞎 | No evidence of microbial cause of inflammation |
| II. Is there a known causal association with the vaccine or vaccination? | | | | | |
| *Vaccine product* | | | | | |
| 1. Is there evidence in published peer reviewed literature that this vaccine may cause such an event if administered correctly? | ⌧ | 🞎 | 🞎 | 🞎 | Several case reports published^2–5^ |
| 2. Is there a biological plausibility that this vaccine could cause such an event? | ⌧ | 🞎 | 🞎 | 🞎 | Exposure to Spike protein antigens |
| 3. In this patient, did a specific test demonstrate the causal role of the vaccine ? | 🞎 | 🞎 | 🞎 | ⌧ | No specific test for this syndrome |
| *Vaccine quality* | | | | | |
| 4. Could the vaccine given to this patient have a quality defect or is substandard or falsified? | 🞎 | ⌧ | 🞎 | 🞎 |  |
| *Immunization error* | | | | | |
| 5. In this patient, was there an error in prescribing or non-adherence to recommendations for use of the vaccine (e.g. use beyond the expiry date, wrong recipient etc.)? | 🞎 | ⌧ | 🞎 | 🞎 |  |
| 6. In this patient, was the vaccine (or diluent) administered in an unsterile manner? | 🞎 | ⌧ | 🞎 | 🞎 |  |
| 7. In this patient, was the vaccine’s physical condition (e.g. colour, turbidity, presence of foreign substances etc.) abnormal when administered? | 🞎 | ⌧ | 🞎 | 🞎 |  |
| 8. When this patient was vaccinated, was there an error in vaccine constitution/ preparation by the vaccinator (e.g. wrong product, wrong diluent, improper mixing, improper syringe filling etc.)? | 🞎 | ⌧ | 🞎 | 🞎 |  |
| 9. In this patient, was there an error in vaccine handling (e.g. a break in the cold chain during transport, storage and/or immunization session etc.)? | 🞎 | ⌧ | 🞎 | 🞎 |  |
| 10. In this patient, was the vaccine administered incorrectly (e.g. wrong dose, site or route of administration; wrong needle size etc.)? | 🞎 | ⌧ | 🞎 | 🞎 |  |
| *Immunization anxiety (Immunization stress related responses - ISRR)* | | | | | |
| 11. In this patient, could this event be a stress response triggered by immunization (e.g. acute stress response, vasovagal reaction, hyperventilation, dissociative neurological symptom reaction etc)? | 🞎 | ⌧ | 🞎 | 🞎 |  |
| II (time): Was the event in section II within the time window of increased risk (i.e. ‘Yes” response to questions from II 1 to II 11 above) | | | | | |
| 12. In this patient, did the event occur within a plausible time window after vaccine administration? | ⌧ | 🞎 | 🞎 | 🞎 |  |
| III. Is there strong evidence against a causal association? | | | | | |
| 1. Is there a body of published evidence (systematic reviews, GACVS reviews, Cochrane reviews etc.) **against** a causal association between the vaccine and the event? | 🞎 | ⌧ | 🞎 | 🞎 | No population based or pharmacovigilance study infirming the risk of hyper-inflammatory syndrome following mRNA COVID-19 vaccines in children |
| IV. Other qualifying factors for classification | | | | | |
| 1. In this patient, did such an event occur in the past after administration of a similar vaccine? | 🞎 | ⌧ | 🞎 | 🞎 |  |
| 2. In this patient, did such an event occur in the past independent of vaccination? | 🞎 | ⌧ | 🞎 | 🞎 |  |
| 3. Could the current event have occurred in this patient without vaccination (background rate)? | 🞎 | ⌧ | 🞎 | 🞎 |  |
| 4. Did this patient have an illness, pre-existing condition or risk factor that could have contributed to the event? | 🞎 | ⌧ | 🞎 | 🞎 |  |
| 5. Was this patient taking any medication prior to the vaccination? | 🞎 | ⌧ | 🞎 | 🞎 |  |
| 6. Was this patient exposed to a potential factor (other than vaccine) prior to the event (e.g. allergen, drug, herbal product etc.)? | 🞎 | ⌧ | 🞎 | 🞎 | Especially, no evidence of previous exposure to SARS-CoV-2 |

Note: Y: Yes; N: No; UK: Unknown; NA: Not applicable.

No

Step 3 (Algorithm) review all steps and ✓ all the appropriate boxes

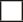

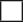

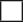

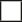

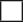

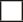

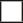

I A. Inconsistent causal association to immunization

III A. Inconsistent causal association to immunization

Yes

Yes

I. Is there strong evidence for other

causes?

II. Is there a known causal association with the vaccine/

vaccination

III. Is there a strong evidence against a causal association?

IV. Review other qualifying factors

Yes

II (Time). Was the event within the time window of increased risk?

Is the event classifiable?

IV D. Unclassifiable

Yes

Yes

II A. Consistent causal association to immunization

IV A. Consistent causal association to immunization

IV B. Indeterminate

IV C. Inconsistent causal association to immunization

Mandatory path

**Notes for Step 3:**

II A: With the available information, it seems likely that the vaccine could be involved in the event (no evidence of other cause, similar cases described in the literature, time window plausible)

IV A: No strong evidence against a causal association, and no previous exposure to a potential factor, i.e., no evidence of SARS-CoV-2 infection despite extensive investigation.

Step 4 (Classification) ✓ all boxes that apply

Adequate information available

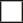

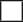

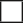

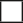

A. Consistent with causal association to immunization

A4. Immunization anxiety-related reaction (ISRR**)

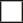

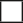

B. Indeterminate

B2. Qualifying factors result in conflicting trends of consistency and inconsistency with causal association to immunization

B1. *Temporal relationship is consistent but there is insuficient definitive evidence for vaccine causing event (may be new vaccine-linked event)

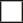

C. Inconsistent with causal association to immunization

C. Coincidental Underlying or emerging

condition(s), or condition(s) caused by exposure to something other than vaccine

A1. Vaccine product-related reaction

(As per published literature)

A2. Vaccine quality defect-related reaction

A3. Immunization error-related reaction

Adequate information not available

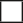

Unclassifiable

Specify the additional information required for classification :

**B1 : Potential signal and maybe considered for investigation*

*** Immunization stress related response*

**Summarize the classification logic in the order of priority:**

With available evidence, we could conclude that the classification is consistent because: there is no evidence of an other cause for the event (especially, no evidence of previous SARS-CoV-2 infection despite extensive investigation), similar cases have been described in the literature, the time window is compatible, there is no strong evidence against a causal association.

Step 1 (Eligibility)

| *Patient ID/Name :* ***Case 6.*** |  | *DoB/Age: 13 years.* |  | *Sex: Male* |
| --- | --- | --- | --- | --- |
| Name one of the vaccines administered before this event |  | What is the Valid Diagnosis? |  | Does the diagnosis meet a case definition? |
| BNT162b2 |  | Hyper-inflammatory syndrome |  | Yes (WHO MIS-C definition) |
| **Create your question on causality here**  Has the BNT162b2 vaccine / vaccination caused Hyper-inflammatory syndrome (The event for review  in step 2 - valid diagnosis) | | | | |
| Is this case eligible for causality assessment? Yes; If, “Yes”, proceed to step 2 | | | | |

Step 2 (Event Checklist) ✓ (check) all boxes that apply

|  | Y | N | UK | NA | Remarks |
| --- | --- | --- | --- | --- | --- |
| I. Is there strong evidence for other causes? | | | | | |
| 1. In this patient, does the medical history, clinical examination and/or investigations, confirm another cause for the event? | 🞎 | ⌧ | 🞎 | 🞎 | No evidence of microbial cause of inflammation |
| II. Is there a known causal association with the vaccine or vaccination? | | | | | |
| *Vaccine product* | | | | | |
| 1. Is there evidence in published peer reviewed literature that this vaccine may cause such an event if administered correctly? | ⌧ | 🞎 | 🞎 | 🞎 | Several case reports published^2–5^ |
| 2. Is there a biological plausibility that this vaccine could cause such an event? | ⌧ | 🞎 | 🞎 | 🞎 | Exposure to Spike protein antigens |
| 3. In this patient, did a specific test demonstrate the causal role of the vaccine ? | 🞎 | 🞎 | 🞎 | ⌧ | No specific test for this syndrome |
| *Vaccine quality* | | | | | |
| 4. Could the vaccine given to this patient have a quality defect or is substandard or falsified? | 🞎 | ⌧ | 🞎 | 🞎 |  |
| *Immunization error* | | | | | |
| 5. In this patient, was there an error in prescribing or non-adherence to recommendations for use of the vaccine (e.g. use beyond the expiry date, wrong recipient etc.)? | 🞎 | ⌧ | 🞎 | 🞎 |  |
| 6. In this patient, was the vaccine (or diluent) administered in an unsterile manner? | 🞎 | ⌧ | 🞎 | 🞎 |  |
| 7. In this patient, was the vaccine’s physical condition (e.g. colour, turbidity, presence of foreign substances etc.) abnormal when administered? | 🞎 | ⌧ | 🞎 | 🞎 |  |
| 8. When this patient was vaccinated, was there an error in vaccine constitution/ preparation by the vaccinator (e.g. wrong product, wrong diluent, improper mixing, improper syringe filling etc.)? | 🞎 | ⌧ | 🞎 | 🞎 |  |
| 9. In this patient, was there an error in vaccine handling (e.g. a break in the cold chain during transport, storage and/or immunization session etc.)? | 🞎 | ⌧ | 🞎 | 🞎 |  |
| 10. In this patient, was the vaccine administered incorrectly (e.g. wrong dose, site or route of administration; wrong needle size etc.)? | 🞎 | ⌧ | 🞎 | 🞎 |  |
| *Immunization anxiety (Immunization stress related responses - ISRR)* | | | | | |
| 11. In this patient, could this event be a stress response triggered by immunization (e.g. acute stress response, vasovagal reaction, hyperventilation, dissociative neurological symptom reaction etc)? | 🞎 | ⌧ | 🞎 | 🞎 |  |
| II (time): Was the event in section II within the time window of increased risk (i.e. ‘Yes” response to questions from II 1 to II 11 above) | | | | | |
| 12. In this patient, did the event occur within a plausible time window after vaccine administration? | ⌧ | 🞎 | 🞎 | 🞎 |  |
| III. Is there strong evidence against a causal association? | | | | | |
| 1. Is there a body of published evidence (systematic reviews, GACVS reviews, Cochrane reviews etc.) **against** a causal association between the vaccine and the event? | 🞎 | ⌧ | 🞎 | 🞎 | No population based or pharmacovigilance study infirming the risk of hyper-inflammatory syndrome following mRNA COVID-19 vaccines in children |
| IV. Other qualifying factors for classification | | | | | |
| 1. In this patient, did such an event occur in the past after administration of a similar vaccine? | 🞎 | ⌧ | 🞎 | 🞎 |  |
| 2. In this patient, did such an event occur in the past independent of vaccination? | 🞎 | ⌧ | 🞎 | 🞎 |  |
| 3. Could the current event have occurred in this patient without vaccination (background rate)? | 🞎 | ⌧ | 🞎 | 🞎 |  |
| 4. Did this patient have an illness, pre-existing condition or risk factor that could have contributed to the event? | 🞎 | ⌧ | 🞎 | 🞎 |  |
| 5. Was this patient taking any medication prior to the vaccination? | 🞎 | ⌧ | 🞎 | 🞎 |  |
| 6. Was this patient exposed to a potential factor (other than vaccine) prior to the event (e.g. allergen, drug, herbal product etc.)? | 🞎 | ⌧ | 🞎 | 🞎 | Especially, no evidence of previous exposure to SARS-CoV-2 |

Note: Y: Yes; N: No; UK: Unknown; NA: Not applicable.

No

Step 3 (Algorithm) review all steps and ✓ all the appropriate boxes

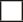

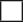

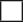

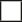

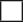

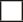

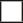

I A. Inconsistent causal association to immunization

III A. Inconsistent causal association to immunization

Yes

Yes

I. Is there strong evidence for other

causes?

II. Is there a known causal association with the vaccine/

vaccination

III. Is there a strong evidence against a causal association?

IV. Review other qualifying factors

Yes

II (Time). Was the event within the time window of increased risk?

Is the event classifiable?

IV D. Unclassifiable

Yes

Yes

II A. Consistent causal association to immunization

IV A. Consistent causal association to immunization

IV B. Indeterminate

IV C. Inconsistent causal association to immunization

Mandatory path

**Notes for Step 3:**

II A: With the available information, it seems likely that the vaccine could be involved in the event (no evidence of other cause, similar cases described in the literature, time window plausible)

IV A: No strong evidence against a causal association, and no previous exposure to a potential factor, i.e., no evidence of SARS-CoV-2 infection despite extensive investigation.

Step 4 (Classification) ✓ all boxes that apply

Adequate information available

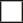

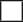

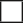

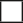

A. Consistent with causal association to immunization

A4. Immunization anxiety-related reaction (ISRR**)

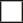

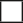

B. Indeterminate

B2. Qualifying factors result in conflicting trends of consistency and inconsistency with causal association to immunization

B1. *Temporal relationship is consistent but there is insuficient definitive evidence for vaccine causing event (may be new vaccine-linked event)

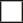

C. Inconsistent with causal association to immunization

C. Coincidental Underlying or emerging

condition(s), or condition(s) caused by exposure to something other than vaccine

A1. Vaccine product-related reaction

(As per published literature)

A2. Vaccine quality defect-related reaction

A3. Immunization error-related reaction

Adequate information not available

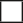

Unclassifiable

Specify the additional information required for classification :

**B1 : Potential signal and maybe considered for investigation*

*** Immunization stress related response*

**Summarize the classification logic in the order of priority:**

With available evidence, we could conclude that the classification is consistent because: there is no evidence of an other cause for the event (especially, no evidence of previous SARS-CoV-2 infection despite extensive investigation), similar cases have been described in the literature, the time window is compatible, there is no strong evidence against a causal association.

Step 1 (Eligibility)

| *Patient ID/Name :* ***Case 7.*** |  | *DoB/Age: 12 years.* |  | *Sex: Female* |
| --- | --- | --- | --- | --- |
| Name one of the vaccines administered before this event |  | What is the Valid Diagnosis? |  | Does the diagnosis meet a case definition? |
| BNT162b2 |  | Hyper-inflammatory syndrome |  | Yes (WHO MIS-C definition) |
| **Create your question on causality here**  Has the BNT162b2 vaccine / vaccination caused Hyper-inflammatory syndrome (The event for review  in step 2 - valid diagnosis) | | | | |
| Is this case eligible for causality assessment? Yes; If, “Yes”, proceed to step 2 | | | | |

Step 2 (Event Checklist) ✓ (check) all boxes that apply

|  | Y | N | UK | NA | Remarks |
| --- | --- | --- | --- | --- | --- |
| I. Is there strong evidence for other causes? | | | | | |
| 1. In this patient, does the medical history, clinical examination and/or investigations, confirm another cause for the event? | 🞎 | ⌧ | 🞎 | 🞎 | No evidence of microbial cause of inflammation, no evidence of leukemia relapse |
| II. Is there a known causal association with the vaccine or vaccination? | | | | | |
| *Vaccine product* | | | | | |
| 1. Is there evidence in published peer reviewed literature that this vaccine may cause such an event if administered correctly? | ⌧ | 🞎 | 🞎 | 🞎 | Several case reports published^2–5^ |
| 2. Is there a biological plausibility that this vaccine could cause such an event? | ⌧ | 🞎 | 🞎 | 🞎 | Exposure to Spike protein antigens |
| 3. In this patient, did a specific test demonstrate the causal role of the vaccine ? | 🞎 | 🞎 | 🞎 | ⌧ | No specific test for this syndrome |
| *Vaccine quality* | | | | | |
| 4. Could the vaccine given to this patient have a quality defect or is substandard or falsified? | 🞎 | ⌧ | 🞎 | 🞎 |  |
| *Immunization error* | | | | | |
| 5. In this patient, was there an error in prescribing or non-adherence to recommendations for use of the vaccine (e.g. use beyond the expiry date, wrong recipient etc.)? | 🞎 | ⌧ | 🞎 | 🞎 |  |
| 6. In this patient, was the vaccine (or diluent) administered in an unsterile manner? | 🞎 | ⌧ | 🞎 | 🞎 |  |
| 7. In this patient, was the vaccine’s physical condition (e.g. colour, turbidity, presence of foreign substances etc.) abnormal when administered? | 🞎 | ⌧ | 🞎 | 🞎 |  |
| 8. When this patient was vaccinated, was there an error in vaccine constitution/ preparation by the vaccinator (e.g. wrong product, wrong diluent, improper mixing, improper syringe filling etc.)? | 🞎 | ⌧ | 🞎 | 🞎 |  |
| 9. In this patient, was there an error in vaccine handling (e.g. a break in the cold chain during transport, storage and/or immunization session etc.)? | 🞎 | ⌧ | 🞎 | 🞎 |  |
| 10. In this patient, was the vaccine administered incorrectly (e.g. wrong dose, site or route of administration; wrong needle size etc.)? | 🞎 | ⌧ | 🞎 | 🞎 |  |
| *Immunization anxiety (Immunization stress related responses - ISRR)* | | | | | |
| 11. In this patient, could this event be a stress response triggered by immunization (e.g. acute stress response, vasovagal reaction, hyperventilation, dissociative neurological symptom reaction etc)? | 🞎 | ⌧ | 🞎 | 🞎 |  |
| II (time): Was the event in section II within the time window of increased risk (i.e. ‘Yes” response to questions from II 1 to II 11 above) | | | | | |
| 12. In this patient, did the event occur within a plausible time window after vaccine administration? | ⌧ | 🞎 | 🞎 | 🞎 |  |
| III. Is there strong evidence against a causal association? | | | | | |
| 1. Is there a body of published evidence (systematic reviews, GACVS reviews, Cochrane reviews etc.) **against** a causal association between the vaccine and the event? | 🞎 | ⌧ | 🞎 | 🞎 | No population based or pharmacovigilance study infirming the risk of hyper-inflammatory syndrome following mRNA COVID-19 vaccines in children |
| IV. Other qualifying factors for classification | | | | | |
| 1. In this patient, did such an event occur in the past after administration of a similar vaccine? | 🞎 | ⌧ | 🞎 | 🞎 |  |
| 2. In this patient, did such an event occur in the past independent of vaccination? | 🞎 | ⌧ | 🞎 | 🞎 |  |
| 3. Could the current event have occurred in this patient without vaccination (background rate)? | ⌧ | 🞎 | 🞎 | 🞎 | Possible macrophagic activation syndrome with hyper-inflammatory syndrome in the context of past leukemia |
| 4. Did this patient have an illness, pre-existing condition or risk factor that could have contributed to the event? | ⌧ | 🞎 | 🞎 | 🞎 | Past leukemia, and possible role of previous chemotherapies that cannot be excluded |
| 5. Was this patient taking any medication prior to the vaccination? | 🞎 | 🞎 | ⌧ | 🞎 | No current treatment, but possible role of previous chemotherapies and immunosuppressive drugs that cannot be excluded |
| 6. Was this patient exposed to a potential factor (other than vaccine) prior to the event (e.g. allergen, drug, herbal product etc.)? | 🞎 | 🞎 | ⌧ | 🞎 | Evidence of previous exposure to SARS-CoV-2 not fully evaluable due to the comorbidity (past leukemia) that may alter immune response. Thus SARS-CoV-2 serology (anti-Nucleocapsid and anti-Spike) may not be fully interpretable. |

Note: Y: Yes; N: No; UK: Unknown; NA: Not applicable.

No

Step 3 (Algorithm) review all steps and ✓ all the appropriate boxes

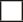

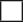

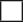

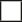

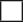

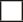

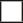

I A. Inconsistent causal association to immunization

III A. Inconsistent causal association to immunization

Yes

Yes

I. Is there strong evidence for other

causes?

II. Is there a known causal association with the vaccine/

vaccination

III. Is there a strong evidence against a causal association?

IV. Review other qualifying factors

Yes

II (Time). Was the event within the time window of increased risk?

Is the event classifiable?

IV D. Unclassifiable

Yes

Yes

II A. Consistent causal association to immunization

IV A. Consistent causal association to immunization

IV B. Indeterminate

IV C. Inconsistent causal association to immunization

Mandatory path

**Notes for Step 3:**

IV B: With the available information, it seems that the potential role of other causes can not be fully excluded (due to a comorbidity, the evidence of previous SARS-CoV-2 could not be fully evaluated, with anti-Nucleocapsid serology negative but not fully interpretable in a context of probable altered immune response). Furthermore, the role of the potential acquired immunodeficiency due to the comorbidity could not be formally excluded

Step 4 (Classification) ✓ all boxes that apply

Adequate information available

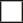

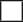

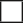

A. Consistent with causal association to immunization

A4. Immunization anxiety-related reaction (ISRR**)

B. Indeterminate

B2. Qualifying factors result in conflicting trends of consistency and inconsistency with causal association to immunization

B1. *Temporal relationship is consistent but there is insuficient definitive evidence for vaccine causing event (may be new vaccine-linked event)

C. Inconsistent with causal association to immunization

C. Coincidental Underlying or emerging

condition(s), or condition(s) caused by exposure to something other than vaccine

A1. Vaccine product-related reaction

(As per published literature)

A2. Vaccine quality defect-related reaction

A3. Immunization error-related reaction

Adequate information not available

Unclassifiable

Specify the additional information required for classification :

**B1 : Potential signal and maybe considered for investigation*

*** Immunization stress related response*

**Summarize the classification logic in the order of priority:**

With available evidence, we could conclude that the classification is Indeterminate because: there is no evidence of an other cause for the event but the evidence of previous SARS-CoV-2 could not be fully evaluated due to a comorbidity, with anti-Nucleocapsid serology negative but not fully interpretable in a context of probable altered immune response.

Step 1 (Eligibility)

| *Patient ID/Name :* ***Case 8.*** |  | *DoB/Age: 12 years.* |  | *Sex: Male* |
| --- | --- | --- | --- | --- |
| Name one of the vaccines administered before this event |  | What is the Valid Diagnosis? |  | Does the diagnosis meet a case definition? |
| BNT162b2 |  | Hyper-inflammatory syndrome |  | Yes (WHO MIS-C definition) |
| **Create your question on causality here**  Has the BNT162b2 vaccine / vaccination caused Hyper-inflammatory syndrome (The event for review  in step 2 - valid diagnosis) | | | | |
| Is this case eligible for causality assessment? Yes; If, “Yes”, proceed to step 2 | | | | |

Step 2 (Event Checklist) ✓ (check) all boxes that apply

|  | Y | N | UK | NA | Remarks |
| --- | --- | --- | --- | --- | --- |
| I. Is there strong evidence for other causes? | | | | | |
| 1. In this patient, does the medical history, clinical examination and/or investigations, confirm another cause for the event? | 🞎 | ⌧ | 🞎 | 🞎 | No evidence of microbial cause of inflammation |
| II. Is there a known causal association with the vaccine or vaccination? | | | | | |
| *Vaccine product* | | | | | |
| 1. Is there evidence in published peer reviewed literature that this vaccine may cause such an event if administered correctly? | ⌧ | 🞎 | 🞎 | 🞎 | Several case reports published^2–5^ |
| 2. Is there a biological plausibility that this vaccine could cause such an event? | ⌧ | 🞎 | 🞎 | 🞎 | Exposure to Spike protein antigens |
| 3. In this patient, did a specific test demonstrate the causal role of the vaccine ? | 🞎 | 🞎 | 🞎 | ⌧ | No specific test for this syndrome |
| *Vaccine quality* | | | | | |
| 4. Could the vaccine given to this patient have a quality defect or is substandard or falsified? | 🞎 | ⌧ | 🞎 | 🞎 |  |
| *Immunization error* | | | | | |
| 5. In this patient, was there an error in prescribing or non-adherence to recommendations for use of the vaccine (e.g. use beyond the expiry date, wrong recipient etc.)? | 🞎 | ⌧ | 🞎 | 🞎 |  |
| 6. In this patient, was the vaccine (or diluent) administered in an unsterile manner? | 🞎 | ⌧ | 🞎 | 🞎 |  |
| 7. In this patient, was the vaccine’s physical condition (e.g. colour, turbidity, presence of foreign substances etc.) abnormal when administered? | 🞎 | ⌧ | 🞎 | 🞎 |  |
| 8. When this patient was vaccinated, was there an error in vaccine constitution/ preparation by the vaccinator (e.g. wrong product, wrong diluent, improper mixing, improper syringe filling etc.)? | 🞎 | ⌧ | 🞎 | 🞎 |  |
| 9. In this patient, was there an error in vaccine handling (e.g. a break in the cold chain during transport, storage and/or immunization session etc.)? | 🞎 | ⌧ | 🞎 | 🞎 |  |
| 10. In this patient, was the vaccine administered incorrectly (e.g. wrong dose, site or route of administration; wrong needle size etc.)? | 🞎 | ⌧ | 🞎 | 🞎 |  |
| *Immunization anxiety (Immunization stress related responses - ISRR)* | | | | | |
| 11. In this patient, could this event be a stress response triggered by immunization (e.g. acute stress response, vasovagal reaction, hyperventilation, dissociative neurological symptom reaction etc)? | 🞎 | ⌧ | 🞎 | 🞎 |  |
| II (time): Was the event in section II within the time window of increased risk (i.e. ‘Yes” response to questions from II 1 to II 11 above) | | | | | |
| 12. In this patient, did the event occur within a plausible time window after vaccine administration? | ⌧ | 🞎 | 🞎 | 🞎 |  |
| III. Is there strong evidence against a causal association? | | | | | |
| 1. Is there a body of published evidence (systematic reviews, GACVS reviews, Cochrane reviews etc.) **against** a causal association between the vaccine and the event? | 🞎 | ⌧ | 🞎 | 🞎 | No population based or pharmacovigilance study infirming the risk of hyper-inflammatory syndrome following mRNA COVID-19 vaccines in children |
| IV. Other qualifying factors for classification | | | | | |
| 1. In this patient, did such an event occur in the past after administration of a similar vaccine? | 🞎 | ⌧ | 🞎 | 🞎 |  |
| 2. In this patient, did such an event occur in the past independent of vaccination? | 🞎 | ⌧ | 🞎 | 🞎 |  |
| 3. Could the current event have occurred in this patient without vaccination (background rate)? | 🞎 | ⌧ | 🞎 | 🞎 |  |
| 4. Did this patient have an illness, pre-existing condition or risk factor that could have contributed to the event? | 🞎 | ⌧ | 🞎 | 🞎 |  |
| 5. Was this patient taking any medication prior to the vaccination? | 🞎 | ⌧ | 🞎 | 🞎 |  |
| 6. Was this patient exposed to a potential factor (other than vaccine) prior to the event (e.g. allergen, drug, herbal product etc.)? | 🞎 | 🞎 | ⌧ | 🞎 | Previous exposure to SARS-CoV-2 could not be formally excluded (Anti-nucleocapsid serology at the limit of significance). |

Note: Y: Yes; N: No; UK: Unknown; NA: Not applicable.

No

Step 3 (Algorithm) review all steps and ✓ all the appropriate boxes

I A. Inconsistent causal association to immunization

III A. Inconsistent causal association to immunization

Yes

Yes

I. Is there strong evidence for other

causes?

II. Is there a known causal association with the vaccine/

vaccination

III. Is there a strong evidence against a causal association?

IV. Review other qualifying factors

Yes

II (Time). Was the event within the time window of increased risk?

Is the event classifiable?

IV D. Unclassifiable

Yes

Yes

II A. Consistent causal association to immunization

IV A. Consistent causal association to immunization

IV B. Indeterminate

IV C. Inconsistent causal association to immunization

Mandatory path

**Notes for Step 3:**

IV B: With the available information, we could not conclude that the vaccine could be involved in the event (previous exposure to SARS-CoV-2 could not be formally excluded (Anti-nucleocapsid serology at the limit of significance).

Step 4 (Classification) ✓ all boxes that apply

Adequate information available

A. Consistent with causal association to immunization

A4. Immunization anxiety-related reaction (ISRR**)

B. Indeterminate

B2. Qualifying factors result in conflicting trends of consistency and inconsistency with causal association to immunization

B1. *Temporal relationship is consistent but there is insufficient definitive evidence for vaccine causing event (may be new vaccine-linked event)

C. Inconsistent with causal association to immunization

C. Coincidental Underlying or emerging

condition(s), or condition(s) caused by exposure to something other than vaccine

A1. Vaccine product-related reaction

(As per published literature)

A2. Vaccine quality defect-related reaction

A3. Immunization error-related reaction

Adequate information not available

Unclassifiable

Specify the additional information required for classification :

**B1 : Potential signal and maybe considered for investigation*

*** Immunization stress related response*

**Summarize the classification logic in the order of priority:**

With available evidence, we could conclude that the classification is Indeterminate because: previous exposure to SARS-CoV-2 could not be formally excluded (Anti-nucleocapsid serology at the limit of significance).

Step 1 (Eligibility)

| *Patient ID/Name :* ***Case 9.*** |  | *DoB/Age: 12 years.* |  | *Sex: Male* |
| --- | --- | --- | --- | --- |
| Name one of the vaccines administered before this event |  | What is the Valid Diagnosis? |  | Does the diagnosis meet a case definition? |
| BNT162b2 |  | Hyper-inflammatory syndrome |  | Yes (WHO MIS-C definition) |
| **Create your question on causality here**  Has the BNT162b2 vaccine / vaccination caused Hyper-inflammatory syndrome (The event for review  in step 2 - valid diagnosis) | | | | |
| Is this case eligible for causality assessment? Yes; If, “Yes”, proceed to step 2 | | | | |

Step 2 (Event Checklist) ✓ (check) all boxes that apply

|  | Y | N | UK | NA | Remarks |
| --- | --- | --- | --- | --- | --- |
| I. Is there strong evidence for other causes? | | | | | |
| 1. In this patient, does the medical history, clinical examination and/or investigations, confirm another cause for the event? | ⌧ | 🞎 | 🞎 | 🞎 | Evidence of previous SARS-CoV-2 infection |
| II. Is there a known causal association with the vaccine or vaccination? | | | | | |
| *Vaccine product* | | | | | |
| 1. Is there evidence in published peer reviewed literature that this vaccine may cause such an event if administered correctly? | ⌧ | 🞎 | 🞎 | 🞎 | Several case reports published^2–5^ |
| 2. Is there a biological plausibility that this vaccine could cause such an event? | ⌧ | 🞎 | 🞎 | 🞎 | Exposure to Spike protein antigens |
| 3. In this patient, did a specific test demonstrate the causal role of the vaccine ? | 🞎 | 🞎 | 🞎 | ⌧ | No specific test for this syndrome |
| *Vaccine quality* | | | | | |
| 4. Could the vaccine given to this patient have a quality defect or is substandard or falsified? | 🞎 | ⌧ | 🞎 | 🞎 |  |
| *Immunization error* | | | | | |
| 5. In this patient, was there an error in prescribing or non-adherence to recommendations for use of the vaccine (e.g. use beyond the expiry date, wrong recipient etc.)? | 🞎 | ⌧ | 🞎 | 🞎 |  |
| 6. In this patient, was the vaccine (or diluent) administered in an unsterile manner? | 🞎 | ⌧ | 🞎 | 🞎 |  |
| 7. In this patient, was the vaccine’s physical condition (e.g. colour, turbidity, presence of foreign substances etc.) abnormal when administered? | 🞎 | ⌧ | 🞎 | 🞎 |  |
| 8. When this patient was vaccinated, was there an error in vaccine constitution/ preparation by the vaccinator (e.g. wrong product, wrong diluent, improper mixing, improper syringe filling etc.)? | 🞎 | ⌧ | 🞎 | 🞎 |  |
| 9. In this patient, was there an error in vaccine handling (e.g. a break in the cold chain during transport, storage and/or immunization session etc.)? | 🞎 | ⌧ | 🞎 | 🞎 |  |
| 10. In this patient, was the vaccine administered incorrectly (e.g. wrong dose, site or route of administration; wrong needle size etc.)? | 🞎 | ⌧ | 🞎 | 🞎 |  |
| *Immunization anxiety (Immunization stress related responses - ISRR)* | | | | | |
| 11. In this patient, could this event be a stress response triggered by immunization (e.g. acute stress response, vasovagal reaction, hyperventilation, dissociative neurological symptom reaction etc)? | 🞎 | ⌧ | 🞎 | 🞎 |  |
| II (time): Was the event in section II within the time window of increased risk (i.e. ‘Yes” response to questions from II 1 to II 11 above) | | | | | |
| 12. In this patient, did the event occur within a plausible time window after vaccine administration? | ⌧ | 🞎 | 🞎 | 🞎 |  |
| III. Is there strong evidence against a causal association? | | | | | |
| 1. Is there a body of published evidence (systematic reviews, GACVS reviews, Cochrane reviews etc.) **against** a causal association between the vaccine and the event? | 🞎 | ⌧ | 🞎 | 🞎 | No population based or pharmacovigilance study infirming the risk of hyper-inflammatory syndrome following mRNA COVID-19 vaccines in children |
| IV. Other qualifying factors for classification | | | | | |
| 1. In this patient, did such an event occur in the past after administration of a similar vaccine? | 🞎 | ⌧ | 🞎 | 🞎 |  |
| 2. In this patient, did such an event occur in the past independent of vaccination? | 🞎 | ⌧ | 🞎 | 🞎 |  |
| 3. Could the current event have occurred in this patient without vaccination (background rate)? | 🞎 | ⌧ | 🞎 | 🞎 |  |
| 4. Did this patient have an illness, pre-existing condition or risk factor that could have contributed to the event? | 🞎 | ⌧ | 🞎 | 🞎 |  |
| 5. Was this patient taking any medication prior to the vaccination? | 🞎 | ⌧ | 🞎 | 🞎 |  |
| 6. Was this patient exposed to a potential factor (other than vaccine) prior to the event (e.g. allergen, drug, herbal product etc.)? | ⌧ | 🞎 | 🞎 | 🞎 | Evidence of previous exposure to SARS-CoV-2. |

Note: Y: Yes; N: No; UK: Unknown; NA: Not applicable.

No

Step 3 (Algorithm) review all steps and ✓ all the appropriate boxes

I A. Inconsistent causal association to immunization

III A. Inconsistent causal association to immunization

Yes

Yes

I. Is there strong evidence for other

causes?

II. Is there a known causal association with the vaccine/

vaccination

III. Is there a strong evidence against a causal association?

IV. Review other qualifying factors

Yes

II (Time). Was the event within the time window of increased risk?

Is the event classifiable?

IV D. Unclassifiable

Yes

Yes

II A. Consistent causal association to immunization

IV A. Consistent causal association to immunization

IV B. Indeterminate

IV C. Inconsistent causal association to immunization

Mandatory path

**Notes for Step 3:**

I A: With the available information, it seems that a potential other cause for the event has been identified (previous SARS-CoV-2 infection).

IV B: While there is a potential other cause for the event has been identified (previous SARS-CoV-2 infection), the potential combined role of SARS-CoV-2 exposure and mRNA COVID-19 vaccine exposure cannot be excluded.

Step 4 (Classification) ✓ all boxes that apply

Adequate information available

A. Consistent with causal association to immunization

A4. Immunization anxiety-related reaction (ISRR**)

B. Indeterminate

B2. Qualifying factors result in conflicting trends of consistency and inconsistency with causal association to immunization

B1. *Temporal relationship is consistent but there is insuficient definitive evidence for vaccine causing event (may be new vaccine-linked event)

C. Inconsistent with causal association to immunization

C. Coincidental Underlying or emerging

condition(s), or condition(s) caused by exposure to something other than vaccine

A1. Vaccine product-related reaction

(As per published literature)

A2. Vaccine quality defect-related reaction

A3. Immunization error-related reaction

Adequate information not available

Unclassifiable

Specify the additional information required for classification :

**B1 : Potential signal and maybe considered for investigation*

*** Immunization stress related response*

**Summarize the classification logic in the order of priority:**

With available evidence, we could conclude that the classification is indeterminate because: there is evidence of an other potential cause for the event (evidence of previous SARS-CoV-2 infection), and the potential combined role of SARS-CoV-2 exposure and mRNA COVID-19 vaccine exposure cannot be excluded.

**References**.

1	World Health Organization. Causality assessment of an adverse event following immunization (‎AEFI)‎: user manual for the revised WHO classification, 2nd ed., 2019 update. 2019. https://www.who.int/publications-detail-redirect/causality-assessment-aefi-user-manual-2019 (accessed Jan 4, 2022).

2	Abdelgalil AA, Saeedi FA. Multisystem Inflammatory Syndrome in a 12-Year-old Boy After mRNA-SARS-CoV-2 Vaccination. *Pediatr Infect Dis J* 2021; published online Dec 21. DOI:10.1097/INF.0000000000003442.

3	DeJong J, Sainato R, Forouhar M, Robinson D, Kunz A. Multisystem Inflammatory Syndrome in a Previously Vaccinated Adolescent Female With Sickle Cell Disease. *Pediatr Infect Dis J* 2021; published online Dec 21. DOI:10.1097/INF.0000000000003444.

4	Yalcinkaya R, Oz FN, Polat M, *et al.* A Case of Multisystem Inflammatory Syndrome in a 12-year-old Male After COVID-19 mRNA Vaccine. *Pediatr Infect Dis J* 2021; published online Dec 14. DOI:10.1097/INF.0000000000003432.

5	Danish Medicine Agency. Danish Medicines Agency investigates a case of inflammatory condition reported after COVID-19 vaccination. Danish Medicines Agency. 2021. https://laegemiddelstyrelsen.dk/en/news/2021/danish-medicines-agency-investigates-a-case-of-inflammatory-condition-reported-after-covid-19-vaccination/ (accessed Jan 4, 2022).
